# Fanconi Anemia as a Window into Premalignant Field Cancerization of the Oral Mucosa

**DOI:** 10.64898/2026.06.13.26354719

**Authors:** Tamar Berger, Frank X. Donovan, Yu-Chien Lin, Shivatheja Soma, Francis May, Kinjal Bhadresha, Christine Krieg, Neelam Giri, Lisa J McReynolds, Armando Filie, Zohreh Khavandgar, Denise M Laronde, Martial Guillaud, Sharon A Savage, David I. Kutler, Wayne Crismani, Eunike Velleuer, Rachel Uppgaard, Ursula L. Harper, K. Olivia Alston, James W. Thomas, Arleen D. Auerbach, Settara C. Chandrasekharappa, Agata Smogorzewska

## Abstract

Head and neck squamous cell carcinoma (HNSCC) evolves through stepwise clonal expansion within genetically altered mucosa fields, yet actionable biomarkers remain undefined. Leveraging Fanconi anemia (FA), a cancer predisposition syndrome with extreme HNSCC risk due to defective DNA interstrand crosslink repair, we profiled premalignant changes in the oral cavity using noninvasive brush biopsies. Consistent with our prior demonstration of genomic instability in FA-associated SCCs, we detected pathogenic *TP53* variants in 26% and copy number alterations in 60.5% in clinically normal-appearing oral mucosa of individuals with FA. These subclinical clonal expansions define candidate biomarkers of early clonal evolution amenable to serial sampling for risk stratification and prevention studies. Since FA-associated SCCs share genomic features with sporadic HNSCC, these findings may extend to the broader population. We also identify somatic reversion of a pathogenic FANCB variant, providing evidence of genomic self-correction and suggesting a potential avenue for gene-based cancer prevention in FA.

**Statement of significance:** Oral mucosa of individuals with Fanconi anemia contains frequent abnormal clones creating a premalignant field that increases cancer risk. The noninvasive brush sampling approach allows repeated measurements, ongoing surveillance, and assessment of prophylactic strategies that may be useful in the prevention of cancers in people with FA and in the general population. Somatic reversion of a pathogenic FANC variant may protect the oral mucosa from DNA repair deficiency and premalignant clonal evolution.

## Introduction

Head and Neck Squamous Cell Carcinoma (HNSCC) remains a formidable disease, with poor survival rate and significant post-treatment morbidity (1). The challenge of treating HNSCC is even greater in Fanconi anemia (FA), a genetic disorder characterized by defective DNA repair, particularly in resolving DNA interstrand crosslinks (ICLs) (2–5). The inability to repair these ICLs in FA results in a dramatically increased incidence of HNSCC at a very young age, over 700 times higher than in the general population (6). This is coupled with an aggressive disease course and limited treatment options, as people with FA cannot tolerate platinum-based chemotherapy (6, 7). Our prior studies revealed that FA-related tumorigenesis is driven by structural variants leading to somatic copy number alterations (CNAs) at critical oncogenic and tumor suppressor loci, often in the context of *TP53* mutation or loss (8). Additionally, we demonstrated that FA-pathway deficient keratinocytes exhibit accelerated epithelial-to-mesenchymal transition, heightened inflammatory signaling, and increased tumor growth rates. Notably, our analysis of sporadic HNSCCs suggests that their pathogenesis may stem from the Fanconi anemia DNA repair pathway being overwhelmed by alcohol- and tobacco-derived aldehyde damage, positioning FA as a valuable model for sporadic HNSCC (8).

Beyond cancer susceptibility, FA also leads to congenital abnormalities and early-onset bone marrow failure, often necessitating allogeneic hematopoietic stem cell transplantation (HSCT) (9). While HSCT results in improved survival, it also markedly increases the risk of solid tumors, particularly HNSCC in FA patients. By age 45, the estimated computed cumulative incidence of HNSCC, calculated using an actuarial method that accounts for the competing risk of non-SCC-related death, is approximately 50% in FA patients without HSCT and approaches 100% in those who have undergone HSCT (10, 11).

Given that most FA-associated HNSCCs originate in the oral cavity and are often preceded by dysplastic lesions, current clinical surveillance strategies focus on early detection through visual exams and surgical or brush biopsies of oral visible lesions (12, 13). This approach is complicated by cancers arising quickly without apparent visible precursor lesions, and by the presence of multiple or extensive lesions that require frequent surgical biopsies (14). Consequently, in the past, many FA patients succumb to their disease within two years due to late-stage diagnoses and intolerance to chemotherapy (8, 14).

To address this critical need, we tested the hypothesis that the normal-appearing mucosa of FA patients harbors genetically abnormal premalignant clones containing variants identified in FA-related HNSCC. We established a robust surveillance framework using simple noninvasive oral brushes to quantify genetic changes including *TP53* variants and genome-wide CNAs in the normal-appearing mucosa. We confirmed the somatic origin of variants by analyzing multiple oral samples per participant and comparing them to their germline DNA. We compared findings from samples from FA patients with those from healthy non-FA volunteers of similar age and sex, as well as older non-FA volunteers. Our analysis revealed somatic variants in non-lesional oral mucosa of individuals with FA, which were absent in healthy volunteers of the same age range and in all but one of the older volunteers. This noninvasive approach will potentially allow for repeated measurements, ongoing surveillance, and assessment of prophylactic strategies. FA presents an ideal model for developing preventive measures to reduce and delay HNSCC onset, with implications extending beyond FA to other high-risk populations, including individuals with a history of HNSCC, HSCT and solid organ transplant recipients, and those at risk due to environmental exposures.

## Results

### Cohort characteristics

Thirty-eight FA patients and 38 non-FA controls consented to our study between June 2023 and December 2024. Two age-based control groups of volunteers without FA and without a history of heavy smoking were included: a younger cohort of 15 volunteers under the age of 50 years, who were close in age to the FA cohort, and an older cohort consisting of 20 participants over the age of 50 years. Median ages of the FA and younger non-FA control cohorts were 28 years (range 12-50) and 35 years (range 23-48), respectively, and did not differ statistically (p=0.53) (**Table 1)**. The third cohort consisting of older non-FA controls was significantly older than the other two cohorts (p<0.0001), with a median age of 64 years (range 51-75). Three controls with a history of heavy smoking (≥15 pack-years [PY]) were excluded (see methods).

**Table 1.** Study Participant Characteristics.

|  | **Fanconi anemia cohort (n=38)** | **Non-FA younger control cohort (n=15)** | **Non-FA older control cohort (n=20)** | **P value\*** |
| --- | --- | --- | --- | --- |
| **Median age at enrollment, years (range)** | 28 (12.6-50) | 35 (23-48) | 64 (51-75.6) | **<0.0001\*\*** |
| **Gender – female, n (%)** | 18 (47) | 9 (60) | 9 (45) | 0.674 |
| **Tobacco history** |  |  |  |  |
| Past or current tobacco smokers, n (%)† | 1 (2.8) | 5 (33.3) | 4 (20) | **0.0062** |
| Median pack years for smokers (range) | Not reported | 2 (0.5-4.25) | 8.5 (1-39) | 0.1761 |
| **Alcohol consumption** |  |  |  |  |
| Active alcohol consumption (%)† | 8 (22.8) | 11 (73.3) | 9 (45) | **0.0033** |
| Average # of beverages per month in active alcohol consumers, median (range) | 2 (0.5-10) | 4 (1-20) | 2 (0.5-28) | **0.0016** |
| **Post HSCT, n (%)** | 25 (65.8) | 0 | 0 | **<0.0001** |
| **Past diagnosis with HNSCC, n (%)** | 7 (18.9) | 0 | 0 | **0.0279** |

The p-value for statistical significance (<0.05) is shown in bold.
\* A significant P value indicates that at least one of the three groups differs significantly from the others.
\*\* There was no difference in age between the FA cohort and younger non-FA control cohort (p=0.53); the older non-FA control cohort was significantly older compared with the other two cohorts (p<0.0001).
† Three FA patients did not respond
HSCT: Hematopoietic stem cell transplantation; HNSCC: head and neck squamous cell carcinoma.

Sex distribution was similar among the three cohorts, with females comprising 47%, 60% and 45% in the FA and in the younger and older non-FA control cohorts, respectively (p=0.674). Tobacco smoking and alcohol consumption were significantly more common among each of the control cohorts compared with the FA cohort. The prevalence of past or current tobacco smokers was 33% and 20% in the younger and older control groups, respectively, versus 2.8% in the FA group (p=0.0062). Similarly, any level of current alcohol consumption was reported in 73% and 45% of the younger and older control groups, respectively, compared with 22.8% in the FA group (p=0.0033). The non-FA control group consisted of healthy adults with no current or past medical problems (n=12) and adults with common chronic conditions, such as anxiety or depression (n=5), benign prostatic hyperplasia (n=3), hypertension (n=3), allergies (n=3), hypercholesterolemia (n=2), arthritis (n=2), diabetes (n=1), coronary artery disease (n=1), Graves’ disease (n=1), ADHD (n=1), obstructive sleep apnea (n=1) and asthma (n=1). Non-FA controls reported no active malignancy or history of malignancy, apart from two participants: one with a previously resected premalignant cervical lesion and one with a history of resected skin cancer at the age of 58.

Most participants with Fanconi anemia were diagnosed with FA during childhood, with a median age at diagnosis of 6.9 years (range 0-46.1). Pathogenic variants in *FANCA gene* caused FA in most participants (n=18, 47.4%). Other affected genes included *FANCD2* (n=5), *FANCC* (n=4), *FANCD1 (BRCA2)* (n=2), *FANCG* (n=2), and *FANCB, FANCI*, *FANCL* (with n=1, each) and causative gene was not reported for four participants. Of the 36 FA patients with available data, eight patients (22.2%) had a history of myelodysplastic syndrome or acute myeloid leukemia. Twenty-five patients (65.8%) had undergone a HSCT: 24 allogeneic HSCT and one autologous HSCT for acute promyelocytic leukemia prior to receiving a diagnosis of FA. Among the 21 allogeneic HSCT recipients who answered the question about presence of graft-versus-host disease (GVHD), nine reported a history of GVHD. Of the 37 FA patients with available data, eleven FA patients (31.4%) had a history of solid cancers, consisting of prior HNSCC (n=7, 18.9%), non-melanomatous skin cancer (n=3), melanoma (n=2), breast cancer (n=3), liver cancer (n=2), vulvar SCC (n=2), thyroid cancer (n=1) and multiple premalignant lesions (n=11).

### *TP53* and copy number alterations are frequent in oral brush samples from patients with Fanconi anemia, but not in controls

To assess and identify genetically abnormal clones in the oral mucosa of FA patients using oral brush biopsy and saliva samples, we recruited patients with FA and two age groups of non-FA healthy controls. We collected oral keratinocytes from six predefined sites of normal-appearing mucosa using noninvasive oral brushes, along with a saliva sample (**Figure 1A**). Targeted sequencing of the *TP53* gene included all exonic regions and flanking intronic sequences to identify single nucleotide variants (SNVs) and insertion-deletions. CNAs were detected using genome-wide single nucleotide polymorphism (SNP) arrays. Any lesion or abnormal appearing site was brushed for cytology, DNA ploidy and molecular analyses. The somatic origin of variants was confirmed by comparing, within each patient, the results of brush biopsies from different oral sites and, when available, the patient’s germline DNA from blood or skin fibroblasts.

**Figure 1.**
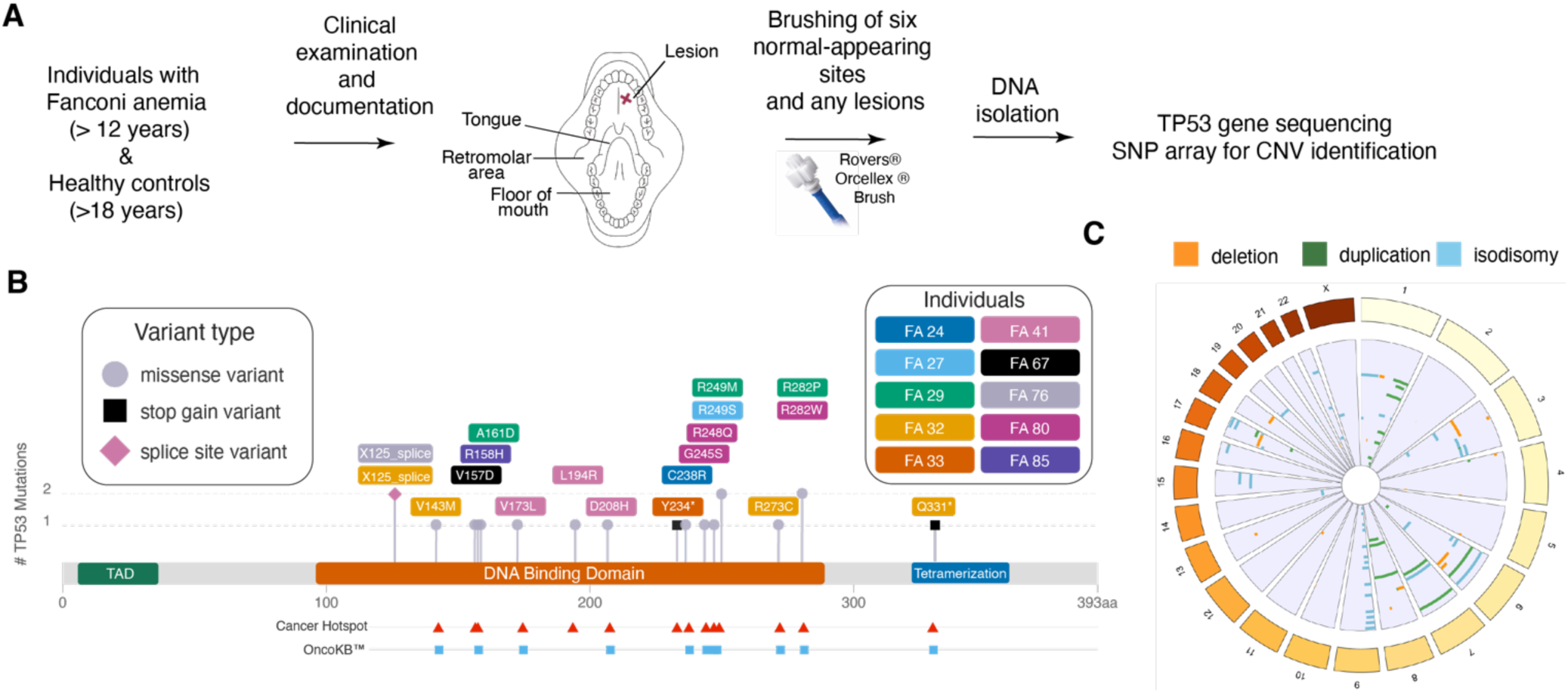
Study’s workflow and variant mapping. **(A)** Graphic representation of the workflow. Six normal-appearing mucosal sites and lesions were brushed using Rovers® Orcellex® brush and isolated DNA was analyzed to identify molecular alterations. **(B)** The diagram mapping nineteen distinct *TP53* SNVs identified in 10 individuals with FA. Each patient is represented by a differently colored rectangle, and variant type (missense, stop gain or splice-site). Diagram was created using MutationMapper (cBioPortal, https://www.cbioportal.org/visualize), with alignment to oncogenic regions via Cancer Hotspots (https://www.cancerhotspots.org/#/home). **(C)** Circos plot showing copy number alterations (CNAs) in normal-appearing mucosa across chromosomes. Each ring corresponds to one individual and displays variants detected in that individual. Data from 23 individuals who harbored CNAs are shown.

A total of 222 oral brush samples collected from non-lesional mucosa had sufficient DNA quantity and quality for *TP53* sequencing in the FA cohort (**Table 2**). Remarkably, eighteen of these samples were positive for 19 distinct somatic *TP53* SNVs (**Figure 1B**, **Table 3**). Somatic *TP53* SNVs were detected in 10 out of the 38 FA individuals (26.3%) at or above 1% variant allele frequency, which we set as a cutoff. Ten of the 18 positive samples originated from the tongue. These 19 distinct clinically significant somatic *TP53* SNVs consisted of 15 missense (nonsynonymous) SNVs, two stop-gain variants and two splice site variants. Four patients harbored multiple distinct *TP53* variants across different brushed oral sites. All variants were pathogenic or likely pathogenic (P/LP) based on the following criteria: absent or negligible frequency in the general population, *in-silico* prediction algorithms and available ClinVar submissions. Eighteen of 19 variants were in the DNA binding domain, and one was in the tetramerization domain of the gene. The majority of the identified somatic missense variants (10/15) fell into high-risk *TP53* variant category, as defined by Poeta *et al.* and EAp53 scoring, which are associated with the poorest survival and the shortest time to the development of distant metastases in non-HPV associated sporadic HNSCC(15, 16).

**Table 2.** Molecular findings in brushed normal-appearing mucosa samples, by cohort.

|  | **FA cohort**<br>**(n=38)** | **Younger control**<br>**cohort (n=15)** | **Older control**<br>**cohort (n=20)** | **P value** |
| --- | --- | --- | --- | --- |
|  | Results from normal appearing oral mucosa samples |  |  |  |
| Participants with a molecular biomarker, n (%) | 23 (60.5%) | 0 | 1 (5) | **<0.0001** |
|  | *TP53* sequencing (allele frequency cutoff of 1%) |  |  |  |
| Number of brush samples with sequencing data | 222 | 81 | 113 | N/A |
| Number of clinically significant somatic variants found, n (%) | 19 (8.5%) | 0 | 2 (1.7) | 0.0891 |
| Participants with a somatic variant, n (%) | 10 (26%) | 0 | 1 (5) | **0.0202** |
|  | Copy number alterations (CNAs) |  |  |  |
| Number of brush samples with SNP array genotyping data | 177 | 87 | 118 | N/A |
| Number of samples with a somatic CNA, n (%) | 62 (35%) | 0 | 0 | **0.0003** |
| Participants with a somatic CNA, n (%) | 23 (60.5%) | 0 | 0 | **<0.0001** |

The p-value for statistical significance (<0.05) is shown in bold.
\* A significant P value indicates that at least one of the three groups differs significantly from the others.
CNA: copy number alterations

**Table 3.**
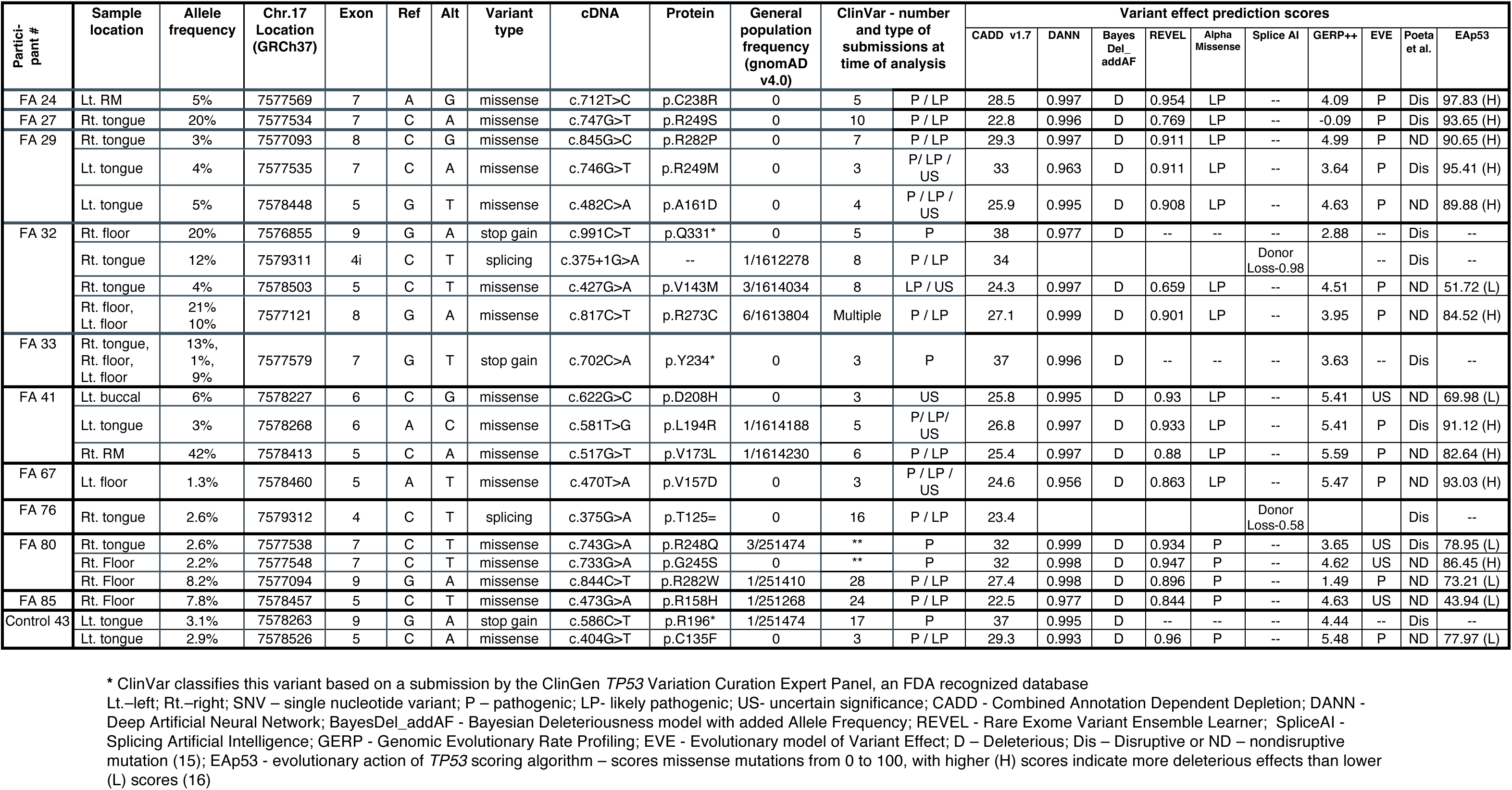
Nineteen *TP53* variants identified in various oral brushes samples from non-lesional mucosa of 10 Fanconi anemia patients and two *TP53* variants identified in a single non-FA control, and their corresponding annotation. Table describes the oral location of variants found, variants’ allele frequency, location on chromosome 17 (Chr.17), whether it is an exonic change (and the exon number) or a splicing alteration, the nucleotide in the reference sequence (Ref) and the alteration found (Alt), the type of functional genetic variants, the notation of the specific genetic variant in the complementary DNA (cDNA) and the protein level change results from the genetic variant in the gene, the frequency of the variants in the general population according to gnomAD v.4, its classifications in ClinVar with number of submissions and in silico prediction scores of variant’s effect using various computational algorithms

In contrast, no somatic *TP53* variants were detected in the normal-appearing oral mucosa in 81 brush samples sequenced of the younger non-FA control cohort (n=15). In the older non-FA control cohort (n=20), only one sample in a single individual had a *TP53* variant, out of the 113 samples sequenced (**Table 2**). The single older non-FA control exhibited two distinct pathogenic *TP53* variants of relatively low frequency (**Table 3**). This was a male in his 60s without oral lesions who denied tobacco smoking or alcohol consumption beyond social use as a young adult, 40 years prior to study enrollment.

At least one CNA was detected through SNP array genotyping of samples from non-lesional oral mucosa in 23 out of 38 participants with FA (60.5%), with numerous samples showing multiple CNAs (**Table 2**, **Figure 1C)**. Of the 177 samples with adequate DNA quantities and high-quality genotype data, 62 samples (35%) were positive for at least one CNA. The most frequently observed CNA was chromosome 9p isodisomy, where tumor suppressors *CDKN2A* (which encodes the two proteins p16 and p14) and *CDKN2B* (which encodes p15) are located. This chromosome 9p aberration was observed in 13/38 FA patients (34.2%). The complete list of CNAs found in the FA cohort appear in **Supplementary Table 1**. Notably, CNAs were not detected in any of the non-FA controls, in over 200 samples genotyped.

In summary, at least one somatic molecular alteration, either a somatic *TP53* variant and/or a CNA, was detected in 23/38 participants with FA (60.5%), while the non-FA controls exhibited zero somatic CNAs and a comparatively low incidence (1/35, 2.8%) of somatic *TP53* variation (p< 0.0001).

### Older age, longer time since transplantation, and prior HNSCC were associated with the presence of molecular variants in Fanconi anemia

We next assessed factors associated with the presence of molecular variants in participants with FA (**Table 4**). Prior HNSCC, older age, and longer time since transplant were all significantly associated with the presence of a molecular variant (p=0.0279, p=0.0247, and p=0.0014, respectively), consistent with known HNSCC risk factors. At least one molecular variant was found in all seven FA participants with a prior history of a HNSCC. A history of prior HSCT was not significantly associated with the presence of a molecular variant, although a possible trend was observed (p = 0.0794), while a history of GVHD was not associated with the presence of a molecular variant (p=1): among 21 allogeneic HSCT recipients with available GVHD data, 9 had a history of GVHD (6 had identifiable variant and 3 did not), and 12 reported no GVHD (8 had identifiable variant and 4 did not). In a multivariable logistic regression analysis including gender, age at time of enrollment and a history of prior HSCT - older age at enrollment was independently associated with higher odds of a molecular variant presence (OR 1.09 per year, 95% CI 1.00-1.19, p=0.042). Post HSCT status showed a non-significant trend towards higher odds of the presence of a molecular variant (OR=4.3, CI 0.91-20.41, p=0.0666), with post-transplant patients having approximately four-fold higher odds of harboring a molecular variant.

**Table 4.**
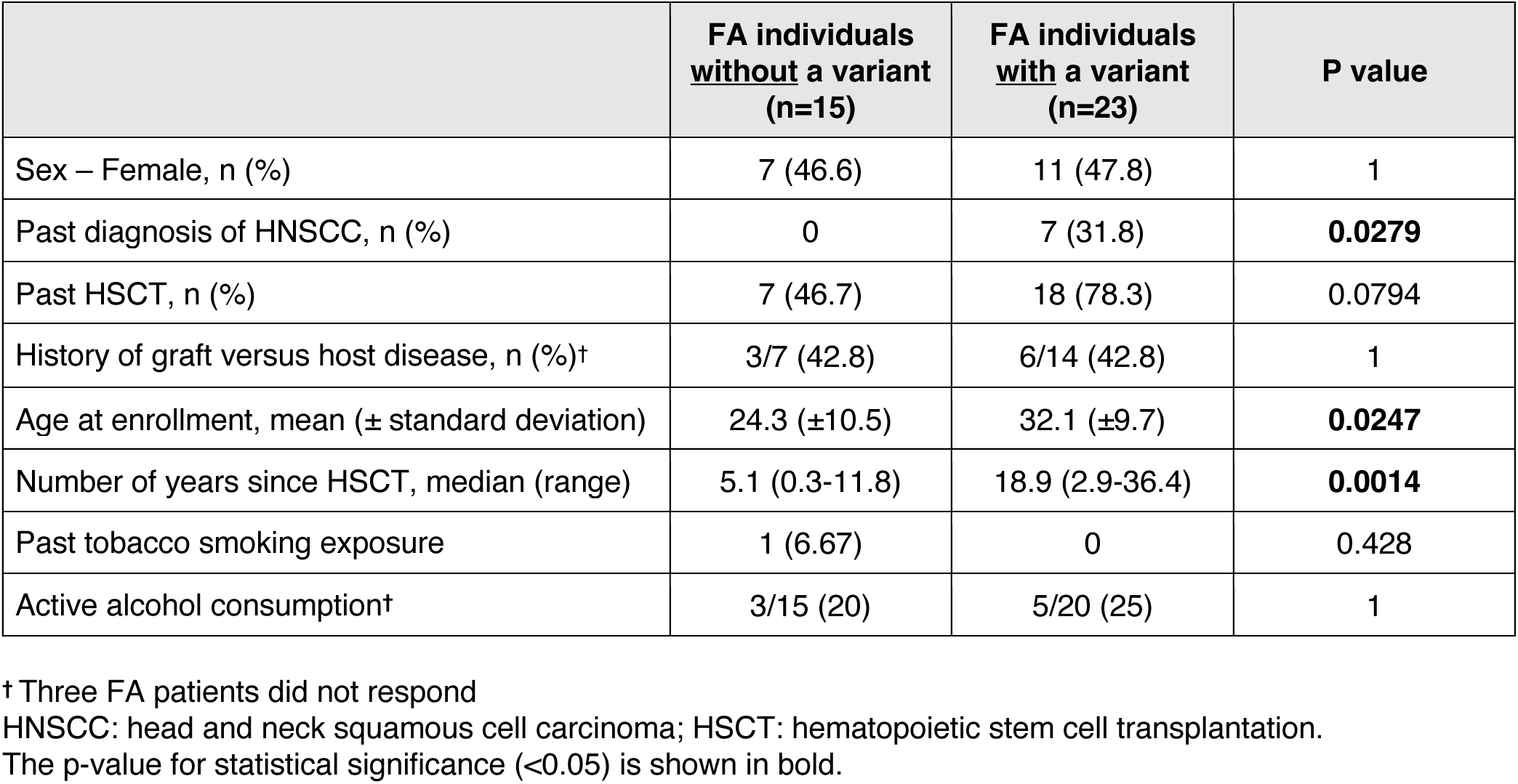
Attributes associated with the presence of molecular variants within the Fanconi anemia cohort.

|  | **FA individuals**<br>**<u>without</u> a variant**<br>**(n=15)** | **FA individuals**<br>**<u>with</u> a variant**<br>**(n=23)** | **P value** |
| --- | --- | --- | --- |
| Sex – Female, n (%) | 7 (46.6) | 11 (47.8) | 1 |
| Past diagnosis of HNSCC, n (%) | 0 | 7 (31.8) | **0.0279** |
| Past HSCT, n (%) | 7 (46.7) | 18 (78.3) | 0.0794 |
| History of graft versus host disease, n (%)† | 3/7 (42.8) | 6/14 (42.8) | 1 |
| Age at enrollment, mean (± standard deviation) | 24.3 (±10.5) | 32.1 (±9.7) | **0.0247** |
| Number of years since HSCT, median (range) | 5.1 (0.3-11.8) | 18.9 (2.9-36.4) | **0.0014** |
| Past tobacco smoking exposure | 1 (6.67) | 0 | 0.428 |
| Active alcohol consumption† | 3/15 (20) | 5/20 (25) | 1 |

† Three FA patients did not respond
HNSCC: head and neck squamous cell carcinoma; HSCT: hematopoietic stem cell transplantation.
The p-value for statistical significance (<0.05) is shown in bold.

A limited number of FA patients reported exposure to tobacco smoking or alcohol consumption, both established risk factors for oral cancer. Eight FA patients reported current alcohol consumption, with an average of 2 beverages per month (median, range 0.5-10). Of those, 5 had identifiable variants - all harbored CNAs, with one patient harboring multiple CNAs and *TP53* variants, and 3 had no variants. Current alcohol consumption at these levels was not associated with variant presence compared with complete abstinence (p=1). The only FA patient who reported past tobacco smoking and current alcohol use had no variants.

Comparison of the three anatomical sites brushed for molecular variants - the retromolar area, lateral tongue and floor of the mouth - did not reveal a regional effect (p=0.1111).

### Suboptimal yield of variant detection from saliva samples

DNA derived from self-collected saliva samples was sequenced from 17 older non-FA controls and 31 FA patients (18 post HSCT). Sequencing of DNA from saliva of 17 older controls revealed no CNAs or *TP53* variants, including in the individual in whom two *TP53* variants (2.9-3.1% allele frequency (AF)) were detected in a brushed biopsy sample, which were not detected in the corresponding saliva sample. Saliva sample sequencing from 31 FA patients, revealed a *TP53* variant in only one patient, despite identification of multiple *TP53* variants in brush biopsies from 10 out of the 31 participants with FA. In the one positive case, the *TP53* variant found in saliva was identical to the one identified in a brush from a lesion, but not in brushes from the normal oral mucosa (**Table 3 and Supplementary Table 2)**. This discrepancy may arise because premalignant lesional cells are more likely to shed into saliva, whereas premalignant cells in the normal-appearing, non-lesional mucosa are still in the basal layer, only sampled by brushing. Alternatively, there may be more premalignant cells in the lesion that increase their detection.

DNA isolated from the saliva from 30 FA patients was analyzed by SNP array. In 10 patients, all post HSCT, the presence of a DNA mixture representing both donor and recipient prevented reliable genotyping and CNA detection. From the 20 patients with reliable SNP array data, saliva samples displayed fewer CNAs than DNA from brushes. Fifteen saliva samples did not exhibit any somatic CNA, despite CNAs being detected in at least one oral brush biopsy in seven of the participants with FA. Five saliva samples were positive for at least one somatic CNA. Two of them shared CNAs with corresponding oral brush samples, two harbored different CNAs from those identified in oral brush samples, and one was observed in a patient with no CNAs detected in oral brush samples (**Supplementary Table 1**). Saliva samples are known to contain many leukocytes (17), which may explain lower sensitivity of saliva analysis across all samples and more likelihood of “contamination” with donor blood.

### Lesions and normal-appearing oral mucosa shared the same *TP53* variants

In addition to brushing the six normal-appearing non-lesional oral mucosal sites, any identified lesion was recorded and brushed for cytology and DNA ploidy analyses and then re-brushed for molecular analysis. Overall, 24 samples from 13 FA individuals had sufficient DNA for sequencing. At least one molecular variant was detected in 13 out of 24 samples (54.2%) (**Supplementary Table 2 and 3**). Six out of 24 lesional samples (25%) collected from four FA individuals, harbored seven distinct clinically significant *TP53* somatic variants, consisting of four missense variants, and three stop-gain variants. In two participants, three of the seven *TP53* variants detected in oral lesions were also present in normal-appearing mucosal sites brushed from the same individual. In one participant, a *TP53* variant present at 9% AF in an ulcerated lesion on the left tongue, was also detected in the normal-appearing mucosa of the same side of the tongue at 3% AF. In another participant, two variants (1% and 12% AF) identified in a lesion on the left lateral tip of the tongue were also evident in normal-appearing floor of the mouth mucosa samples at a higher AF of 10-20%. Cytology of cells collected from these six lesions were read as negative (n=1), atypical squamous cells likely representing reactive changes (n=4), and atypical squamous cells likely representing dysplastic changes with aneuploid (high DNA index) (n=1). The latter lesion was further biopsied, with normal histology reported.

Eighteen of 24 lesional samples collected from 12 FA individuals contained adequate quantity and quality of DNA enabling CNA detection, while six samples contained a DNA mixture of donor cell leukocytes prohibiting genotyping. Among the 18 lesional samples, at least one CNA was detected in 10 samples (55.5%) across 7 FA individuals. Cytology results for these ten samples with detected CNAs were negative (n=5), positive for atypical squamous cells (n=3), suspicious cells (n=1), and dysplastic cells (n=1). Of the latter two samples, one had non-diagnostic DNA-image-cytometry and a surgical biopsy performed excluded malignancy. The second sample showed atypical squamous cells on DNA-image-cytometry indicative of dysplasia with aneuploidy. However, further biopsy showed no premalignant or malignant histology. Eleven lesions had negative cytology and among the nine with available DNA ploidy results, all were diploid, and none exhibited any molecular variants.

### Clinical follow-up

Out of the 38 FA patients recruited to our study, 17 patients had over a year of clinical follow-up. The median follow-up time for the entire FA cohort was 1.03 (range 0.44-2.05) years. During that time, three patients were diagnosed with oral SCC and two were diagnosed with a dysplastic lesion. One patient developed a malignant lesion located at the floor of the mouth, confirmed by cytology, approximately 10 months after it was brushed as a normal-appearing site in our study. This case is presented in **Figure 2** and described in the legend.

**Figure 2.**
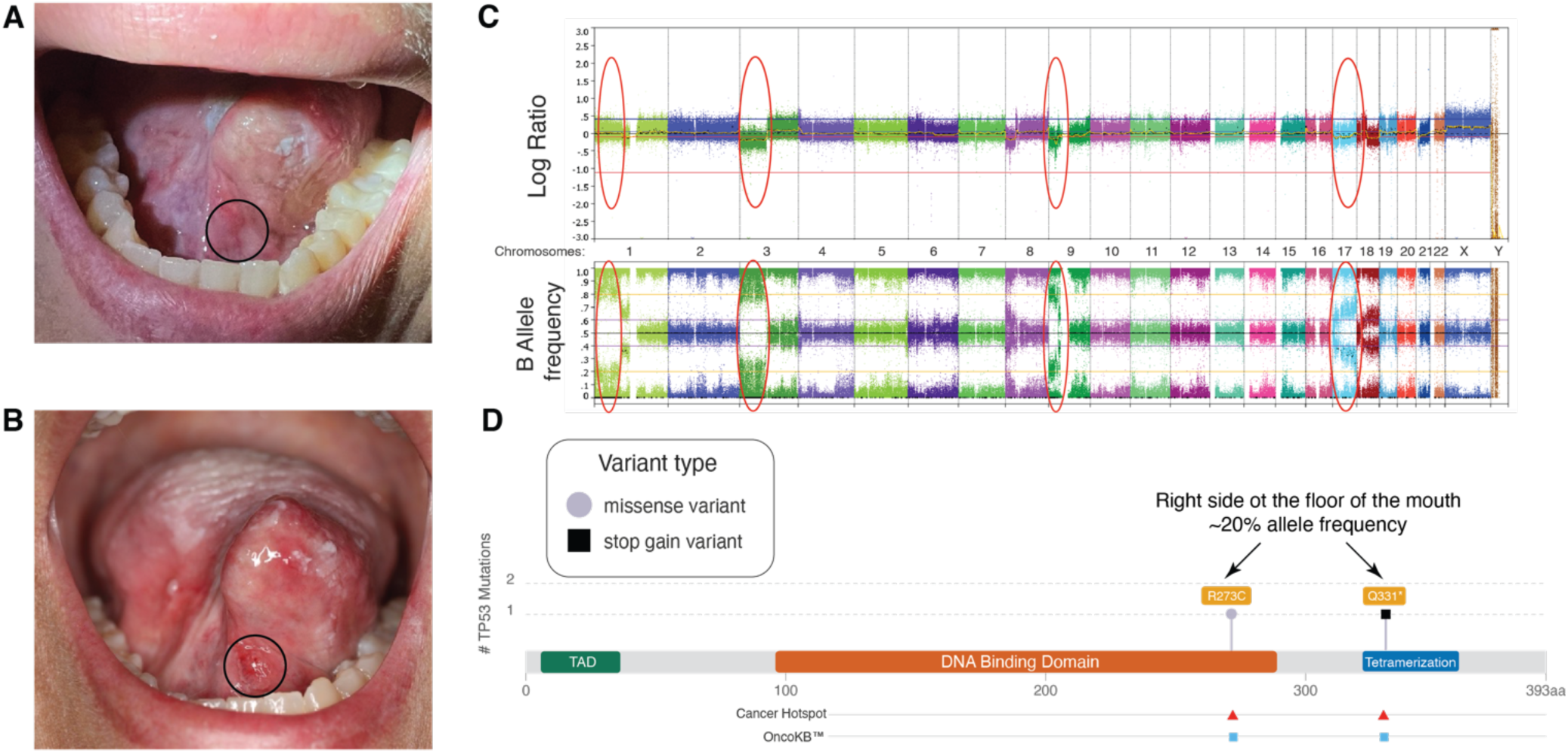
Molecular findings in brush biopsy of non-lesional mucosa at the floor of the mouth/base of tongue of an individual in their 20s with FA. **(A)** Photograph of the floor of the mouth/ventral tongue at first visit. Approximate area of brushing is circled. **(B)** Same area photographed 10 months later with a visible lesion circled. **(C)** SNP array results of brushing from the first visit capturing multiple CNAs across the genome, with some of the loss/isodisomy events circled in red. These include recurrent high-risk events chromosome 9p and 17p loss, affecting tumor suppressors genes *CDKN2A*, *CDKN2B,* and *TP53*, respectively. A complete list of CNAs is listed in **Supplementary Table 3**. **(D)** A diagram of the *TP53* gene, showing location of the two pathogenic somatic variants with 20-21% allele frequency identified from the first visit brushing of the floor of the mouth circled in A. Full description of variants is available in **Supplementary Table 2.**

Another two patients were diagnosed with SCC of the tongue. One had a preceding lesion at the time of brushing, while the second had no lesion and was diagnosed with a dysplastic lesion and cancer approximately 10 months post recruitment and sampling of predefined normal-appearing sites in the mouth. Two other patients were diagnosed with premalignant dysplastic oral lesions during the time of follow-up. In all these patients, both a *TP53* variant and multiple CNAs were identified upon their first brushing.

Of the 35 non-FA controls enrolled in our study, 26 were available for follow-up. They reported no changes in their oral health 12-18 months after enrollment and sampling, including the single older control participant with identified *TP53* variants.

### Reversion of a pathogenic *FANCB* variant in oral mucosal

One of the participants with FA in this study (FA-38) has previously been shown to exhibit somatic mosaicism in peripheral blood. As we previously reported (18), the pathogenic intragenic duplication in the X-linked *FANCB* gene declined from 93% to 8% over an 11-year period through microhomology-mediated recombination. Remarkably, skin fibroblasts from this individual also showed evidence of reversion, with approximately 8% of cells carrying the corrected wild-type allele. We concluded that reversion within the hematopoietic compartment protected this participant from bone marrow failure, as his blood counts remained in the normal range and he has not required HSCT.

Given the high instability of the pathogenic duplication, we hypothesized that somatic reversion might also occur in the oral mucosa and could provide protection against malignant transformation. Clinical examination of the oral cavity revealed no visible abnormalities, and no *TP53* variants or CNAs were detected in any of the brush biopsy samples. To directly assess reversion, DNA isolated from seven independent oral brushings was analyzed for the *FANCB* duplication by digital droplet PCR (ddPCR). The duplication was only detected at low levels (5–10%), whereas fibroblasts analyzed contemporaneously showed approximately 92% mutant allele **(Figure 3).** These findings indicate that somatic reversion of the pathogenic duplication to the wild-type allele occurred in the patient’s oral mucosa.

**Figure 3.**
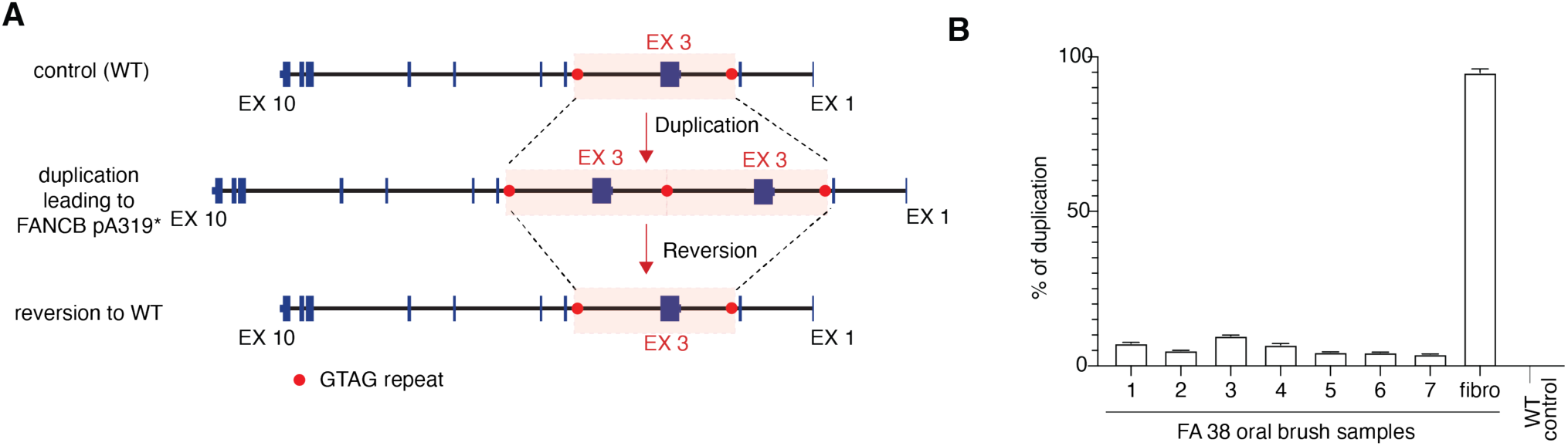
A case of reversion of a *FANCB* mutation in the oral mucosa in a study participant with FA. **(A)** Diagram showing wild type (WT) locus of the *FANCB* gene on the X chromosome (top). Germline duplication on the region highlighted in pink led to formation of a non-functional *FANCB* allele (pA319*) (reference 18). Reversion of the duplication occurs through microhomology-mediated recombination where the abnormal copy is precisely removed from the genome (bottom). **(B)** Assessment of the *FANCB* gene duplication in genomic DNA isolated from oral brushes collected from participant FA 38. Digital droplet PCR (ddPCR) was performed on the six predefined normal-appearing sites (samples 1-6) and the buccal mucosa (sample 7). For comparison, DNA isolated from the fibroblasts of the same participant (fibro) and a control cell line which lacks the duplication were used.

## Discussion

Using targeted *TP53* sequencing and genome-wide SNP array analysis to identify *TP53* SNVs and genome-wide CNAs, respectively, we demonstrated widespread somatic clonal fields in the normal-appearing oral mucosa of patients with FA (26% and 60.5% respectively). Somatic clones were more frequent in patients with FA who were older, had a prior history of HNSCC, and had a longer interval since HSCT. Notably, detectible clones were largely absent in non-FA individuals of similar ages. Only one older non-FA individual harbored *TP53* variants, without CNAs, in a single sample out of nearly 200 control samples sequenced (0.5% and 0% respectively).

These findings suggest that the identified clones represent an intrinsic keratinocyte phenotype in patients with FA. They likely reflect the natural somatic molecular evolution of normal-appearing oral mucosa before the emergence of visible precursor lesions or cancer, providing important insight into early carcinogenic processes in the mucosal squamous cell carcinoma. Although our studies focused on the oral mucosa, it is highly likely that similar clonal mutagenesis is occurring in the mucosa of the upper esophagus, and anogenital region, which are the other sites of frequent SCC formation in FA patients and have a similar genomic alteration landscape (8).

Previous studies of the molecular changes in the mucosa of patients with FA have been limited. The first study to assess the presence of variants in the oral mucosa of FA patients utilizing the oral brush, was limited to non-transplant recipients (19). Only 12 selected microsatellite markers located on four chromosomes: 3p, 9p, 11q, and 17p were analyzed. LOH was detected in 14 out of 141 (9.9%) non-transplanted patients with FA at a median age of 25 (range 6-52). The most frequent LOH occurred at chromosomes 9p (12/141 individuals) and 3p (5/141 individuals). This is consistent with our study, in which 9p LOH was the most frequent copy number event. 9p LOH appeared in a third of the patients with FA studied by us, which is a higher proportion than identified by Smetsers and colleagues(19), most likely due to our inclusion of HSCT recipients and those with past HNSCC.

Another study of FA oral brush samples selected retrospectively from a biobank, included 17 samples from FA patients who later developed oral SCC and compared them to 25 samples from matched FA patients who did not develop oral SCC (20). The presence of SNVs in specific targeted genes and CNAs from oral brush cells was associated with development of oral cancer, although that cancer cohort was enriched for patients who were diagnosed with oral cancer in less than 3 months after brushing, raising the possibility that the brushings already included malignant cells.

Finally, Errazquin et al. conducted a prospective study analyzing saliva samples from 16 patients with FA without clinically diagnosed oral premalignant lesions or SCC at enrollment, using deep sequencing of a selected panel of cancer-associated genes (21). Nine of 16 patients with FA had SNVs in *TP53* with AF as low as 0.07% and in most cases below 1%. Six patients with variants and available follow up data subsequently developed oral precursor lesions or cancer within 23-630 days, whereas patients without variants developed significantly fewer precursor lesions or cancer. However, CNAs were not assessed in that study.

Given the potential for broader implementation afforded by a simpler saliva collection process, we incorporated saliva sampling into our study. However, variant detection from saliva was limited, with a lower detection rate (at an AF of ≥ 1%) compared with oral brush samples. Among 10 FA patients with *TP53* variants in oral brush samples of normal-appearing sites, saliva detected a *TP53* variant in only one patient, matching the patient lesional samples findings. Importantly, while CNAs were detected in oral brush samples, a third of the saliva samples exhibited a DNA mixture in post HSCT patients preventing reliable genotyping and CNA detection. Unlike oral brush samples, saliva contains a higher proportion of blood cells admixed among sloughed oral keratinocytes(17). In HSCT recipients, this results in a mixture of donor derived DNA (from hematopoietic cells) and recipient-derived DNA (from oral keratinocytes) present in saliva (22, 23), thereby causing a dilution effect on somatic variants of recipient-origin, and impeding reliable genotyping and CNA detection. Due to the suboptimal yield of variant detection using saliva samples, we discontinued saliva collection for molecular analysis relying on sampling with brush biopsies.

### Potential sources of DNA damage in the squamous mucosa

Most participants with FA who harbored positive molecular markers reported no known exposure to tobacco or alcohol, major risk factors typically linked to HPV-negative head and neck cancers (1). Moreover, no association was observed between variant presence and patient-reported exposure to these risk factors. This observation points to alternative sources of DNA damage. Endogenous aldehydes, including formaldehyde and other reactive aldehydes, are well-established drivers of bone marrow failure and leukemia (24–32). Building on this knowledge, our group recently demonstrated that formaldehyde and lipid-derived aldehydes are particularly toxic to oral keratinocytes lacking a functional FA pathway (33), underscoring their potential role as endogenous mutagens in the squamous mucosa. Additional contributors may include environmental pollutants, dietary sources of reactive aldehydes, and toxic metabolites produced by the oral microbiome.

Together, these exposures may also play a role in the pathogenesis of sporadic cancers. Based on prior findings that the number of focal somatic copy number alterations in HNSCCs correlates with increased tobacco pack-years exposure in sporadic cancers (8), we suggest that the exogenous toxic aldehydes present in tobacco and alcohol may overwhelm even the intact FA DNA repair pathway and lead to unrepaired DNA damage resulting in CNAs that drive cancer development in the general population (8). Endogenous metabolites would be expected to add to the load of damage over human lifetime even in those without FA.

### Clonal expansion of epithelial cells

Multiple studies uncovered a rich landscape of clonally expanded cancer-driving somatic variants of low allele frequency in aging colorectal, esophageal, skin epithelium and hematopoietic cells (34–43). However, the extent of molecular alterations we observe in FA patients is striking and underscores the critical role of the FA pathway in maintaining genomic integrity of the epithelial mucosa to prevent carcinogenesis.

What determines whether the clones remain constrained or progress towards malignancy is not fully understood, but it is likely influenced by a combination of tissue-specific properties, environmental exposures, and genomic context, including the nature of the initiating mutation(37). In the setting of FA, the clonal expansion may be fueled by multiple forces. We posit that *TP53* variants, provide an increased advantage to cells compromised by the underlying DNA repair deficiency when they are surrounded by FA-deficient cells. Keratinocytes deficient in the FA pathway grow more slowly than the complemented counterparts(33), thus harboring a *TP53* mutation would provide a significant growth advantage to the FA pathway deficient keratinocytes, allowing these cells to substantially expand and replace the *TP53* wildtype cells. Additional CNAs commonly present in individuals with FA may further enhance this selective advantage. In non-FA individuals without the increased burden of DNA damage, the expansion of clones would be limited by no or minimal advantage over the surrounding mucosa. However, in heavy smokers and alcohol drinkers, or those with high environmental exposures, the increased DNA damage load might induce FA-like changes in the mucosa, allowing for substantial clonal expansion when *TP53* is mutated, increasing the likelihood of cancer. Lastly, reduced immune surveillance may contribute to potential clonal expansion in the mucosa of individuals with FA. An immunosuppressive environment was observed in genomically unstable cancers, including FA-associated HNSCCs ((44), Berger and Smogorzewska, unpublished). This is further compounded by the systemic immunosuppressive state of HSCT recipients that contributes to increased risk of solid cancers(45). We hypothesize that DNA damage present in FA mucosa may lead to early immune exhaustion allowing immune evasion and expansion of mutant clones.

In contrast to the growing understanding of age-associated accumulation of SNVs in cancer-related genes within normal tissues, far less is known about how age influences the presence and frequency of CNAs in non-cancerous tissues. Previous studies varied in their power to detect CNAs (36, 37, 39), while in the normal esophagus and colorectal epithelium CNAs were infrequent even in large clones (38). Thus, the prevailing view is that acquired chromosomal instability is confined to cancerous cells or later stages of cancer development. However, genomic instability is a key characteristic of FA. The presence of CNAs harboring clones in the normal oral mucosa of 60% of our FA cohort, including multiple CNAs in many patients, and their complete absence in non-FA controls of any age, reflects oral keratinocyte genomic instability that accumulates with age, over successive cell divisions. It remains to be determined if CNAs might also be present in normal appearing mucosa of non-FA individuals with high environmental exposure.

### Factors associated with molecular changes in the mucosa

In univariate analysis, older age at enrollment and longer time since HSCT were significantly associated with the presence of oral molecular variants in the FA cohort (**Table 4**). In a multivariable analysis, to avoid collinearity we included age at enrollment but excluded time since HSCT. Each additional year of age was associated with a 9% increase in the odds of harboring a molecular variant. Another covariat that was found to be significantly associated with the presence of molecular variants in the normal-appearing mucosa was a history of FA-related HNSCC. Seven FA patients had a history of oral SCC, diagnosed a median of 5 years (range 1-13 years) prior to study enrollment. All of these patients had at least one molecular variant, with six out of the seven patients testing positive for one or more *TP53* SNVs. Unfortunately, tumor genomic data were not available for variant comparison.

In sporadic HNSCC, local cancer recurrence is attributed to the existence of a ‘cancerization field’ of molecularly altered clonal cells remaining within the surgical margins after cancer resection (46). Therefore, we expected to find premalignant clones in FA patients with a history of oral cancer. However, these genetically abnormal clones had diverse *TP53* variants and were identified even years after cancer removal, in locations other than the primary tumor site. This suggests that these findings may be linked not only to the cancerization field effect but to the constant production of new variants throughout life in patients with FA.

Another known risk factor for HNSCC, prior HSCT, showed a near-significant association with variant presence in univariate analysis (p=0.0794), which may reflect limited statistical power. A similar trend was observed in multivariable analysis, with transplant recipients demonstrating an estimated 4.3-fold greater likelihood of a molecular variant (p=0.0666). This finding aligns with prior retrospective natural history cohort studies, which reported that the age-specific hazard of SCC was 4.4-fold higher in patients who had undergone transplantation compared with those who had not (10). Our study had limited power to detect an association between prior GVHD, another well-established risk factor for HNSCC (10, 45, 47–50), and the presence of somatic premalignant clones (p=1), or to assess for the association of variants with past total-body irradiation exposure due to the small number of affected participants studied.

### Implications for reversion of the pathogenic variant in the oral mucosa

In the context of FA, somatic mosaicism refers to the occurrence of a spontaneous reversion of a germline pathogenic variant in a population of cells. This results in two or more genetically distinct populations of cells within the same patient, with a subset of cells regaining functional activity of the FA DNA repair pathway, at times described as “natural gene therapy” (51, 52). The majority of somatic mosaicism has been seen in the hematopoietic compartment, where there is a strong selection for such events. In the gene therapy trial setting, this selective advantage of wild type hemopoietic cells was exploited, resulting in their progressive expansion (53). The reported incidence of spontaneous somatic mosaicism in the hematopoietic compartment varies depending on the diagnostic criteria applied and is estimated at 15-25% of FA patients(51).

Here, we have reported the first case of somatic mosaicism leading to reversion mutation in the oral mucosa of a FA patient (FA 38). This patient has a mild FA hematological phenotype attributed to a known somatic mosaicism in the *FANCB* gene, with reversion of the pathogenic mutation evident in the hematopoietic compartment (18). A high percentage of somatic reversion of the *FANCB* mutation was also observed in the patient’s oral mucosa, suggesting a selective advantage of *FANCB*-proficient keratinocytes. This is consistent with our *in-vitro* observations, where wild type keratinocytes have a strong growth advantage against *FANCA*-deficient keratinocytes (33). We speculate that this spontaneous mutation reversion may potentially protect against the FA DNA repair pathway deficiency phenotype and the premalignant clonal evolution and expansion. These findings support the feasibility of therapeutic correction of the oral mucosa, either through ex vivo gene therapy with subsequent grafting or through direct *in vivo* gene delivery, as a potential strategy for cancer prevention in FA patients.

### Study limitations

Despite the small number of participants with FA in our study, we reached highly statistically and biologically relevant conclusions. However, these studies need to be expanded to further validate our findings and uncover other FA-related factors that are associated with the presence of variants in the normal-appearing oral mucosa that may allow for better clinical risk stratification of patients.

Assessment of lesions was a secondary aim and was constrained by the small number of lesions brushed and DNA yield, as clinical cytology analysis was prioritized over molecular analysis. Consequently, definitive conclusions regarding molecular alterations in lesions could not be drawn. Nevertheless, several noteworthy observations were made, including that the overall frequency of detected variants resembled that of the normal-appearing mucosal samples, and that *TP53* stop-gain variants were more frequent in brushed lesions than in normal-appearing mucosa, where missense variants predominated. Lesional *TP53* variants were detected in four patients, all with a history of prior HNSCC, and one patient developed SCC within one year of follow up. In two out of these four patients, *TP53* variants were also shared in samples from normal-appearing mucosal sites, most probably representing presence of a field of genetic alterations.

### Toward Clinical Translation

In sporadic HNSCC, assessment of prevention treatments effectiveness relies on lesion surveillance and cancer development as a primary outcome (54). However, these outcomes may not provide sufficient lead time for effective intervention in patients with FA due to fast development and aggressive nature of their tumors. There is a critical need for a surrogate biomarker of cancer development in FA patients prior to the emergence of clinical precancerous and cancerous lesions. We believe that the *TP53* variants and CNAs that were assessed in this study, may serve as surrogate biomarkers to evaluate preventative treatment effectiveness in FA-associated oral SCC, incorporated as inclusion criteria and secondary outcomes in primary or secondary prevention trials. Additionally, these biomarkers may support earlier diagnosis of cancer occurrence and recurrence. Ultimately, our goal is to shift the paradigm of HNSCC management in FA patients from reactive treatment to active prevention, using targeted interception strategies to improve survival. Finally, by leveraging FA as a model system, our findings on prevention therapies and diagnostic screening may inform strategies for other high-risk populations predisposed to HNSCC, such as heavy tobacco-smokers, alcohol consumers and other genomic instability syndromes, broadening the impact of this work far beyond FA.

## Methods

### Study participants

This study was conducted at the Rockefeller University Hospital (RUH) and included two age-based non-Fanconi anemia control cohorts and a FA cohort. Adult volunteers aged 18 to 75 years, without FA, or patients with FA above the age of 12 years, were eligible to participate in the study if there was no history of severe bleeding, cognitive impairment or pregnancy. Because we previously demonstrated a correlation between pack-years (PY) of tobacco smoking and the number of focal CNAs in sporadic HNSCC (8), and FA patients are aware that smoking increases the risk of HNSCC, we restricted our analysis to samples from volunteers who were nonsmokers or had accumulated < 15PY of smoking within the past 15 years. Therefore, we excluded two older active smokers (with 24- and 17- PY, each), and one older former smoker (19.5-PY) who had quit smoking in the past 10 years. Non-FA volunteers were recruited through the RUH clinical research office. FA patients were recruited by actively approaching those in the International Fanconi Anemia Registry (IFAR) and during the Fanconi Cancer Foundation (FCF) annual symposium. Written informed consent including photo consent forms were obtained from all participants or their legal guardians prior to sampling. All consent processes were completed through the REDCap system (ver. 14.5.1), a secure web application for building and managing online surveys and databases (55). The study was approved by the Rockefeller University Institutional Review Board (IRB), New York (protocol number TBE-1041).

### Study’s procedures and workflow

Study procedures included targeted physical exam of the oral cavity, pictures and video recording of the oral mucosa of FA patients using a *Karl-Storz* flexible video rhino-laryngoscope (11102CMK), lesion documentation using a mouth map and six oral brush biopsies of normal-appearing mucosa at predefined locations. Any lesion was separately brushed as previously described (19). Blood sampling was done for germline DNA isolation, and a skin punch biopsy was performed only in adult FA participants post HSCT, for the purpose of germline DNA isolation from cultured skin fibroblasts. Saliva collection was added to our protocol at a later stage of the study, thus some of the participants did not have saliva testing. Study’s workflow is depicted in **Figure 1A**.

Most study visits and procedures took place at the Rockefeller University Hospital (RUH). Some of the FA patients’ visits took place at adult meetings of FA patients organized by the Fanconi Cancer Foundation or in participants’ homes. In these cases, only oral cavity examination, noninvasive brush biopsies and saliva samples were collected. To increase the FA cohort sample size, sampling was performed by collaborators from the NIH after remote consent was obtained by the study team. This was approved by the NIH IRB (IRB ID 001109 / CR001091). Demographic details, medical and cancer history, social history including tobacco and alcohol exposure were collected at baseline using REDCap. Participants were followed annually. All participants were asked to report on any new concerning oral findings, and pathology reports were obtained for any incisional or brush biopsies.

To confirm the somatic origin of oral brush findings, any genetic change in oral keratinocytes was compared across different sites brushed within the same subject and variant’s allele frequency and with the subject’s germline DNA extracted from a blood sample or skin biopsy if available. Oral lesions were also analyzed for cytology and DNA ploidy as described below and findings were compared to molecular findings. All molecular findings were correlated back with the clinical documented exam performed prior to obtaining the samples.

### Blood samples

In healthy individuals and participants with FA with no history of HSCT, germline DNA was isolated from peripheral leukocytes using Zymo Research Quick-DNA Microprep kit and quantified with Invitrogen Qubit® dsDNA BR Kit.

### Skin biopsy

A 4mm skin punch biopsy was performed to derive cultured skin fibroblasts for germline DNA analysis in adult FA patients who have undergone prior HSCT and without available preexisting germline genomic data for comparison. All samples were coded.

### Oral brush biopsies and saliva collection

Orcellex® (Rovers Medical Devices) brushes were used to sample six predefined oral locations of normal-appearing mucosa that are known to be prone to SCC development: bilateral retromolar area, lateral tongue and floor of the mouth (**Figure 1**). According to former protocols (19), one or two brushes were rotated at least ten times at each of the locations. Any visible lesions were brushed separately for cytology, DNA ploidy, and molecular testing. Samples were placed in BD SurePath™ preservation solution. Saliva samples were collected using DNAgenotek Oragene DISCOVER (OGR-600) collection kits following the company instructions. Variant detection yield was compared between these two noninvasive methods of sampling - oral brush biopsies and saliva collection.

### DNA isolation

Oral keratinocytes collected through oral brushing of normal-appearing mucosa and lesions were subjected to molecular testing. To facilitate storage, shipment and sharing of additional samples from remote locations, we stored oral brush samples in ‘BD SurePath’, a widely used ethanol-based cytology preservative. Although not ideal for subsequent cell lysis, after optimizing our DNA isolation protocols, we obtained satisfactory high-molecular-weight DNA from samples stored up to 77 days, with no consistent DNA degradation or base-level alterations (see **Supplemental methods**).

Brush biopsies samples were processed using Qiagen DNA isolation kit optimized protocol. DNA from saliva samples was isolated using DNAgenotek prepIT-L2P commercially available kit. DNA quantification was done with Invitrogen Qubit® dsDNA kit and nanodrop assay. Ethidium bromide (EtBr) gel electrophoresis was used to determine the molecular weight of selected DNA samples and to exclude fragmentation.

### TP53 sequencing

*TP53* was sequenced on the Illumina Miseq platform, using the Paragon CleanPlex *TP53* kit (Paragon Genomics, Inc), to identify single nucleotide variants (SNVs) and insertion-deletion variants (indels). The kit targets all exonic regions and flanking intronic sequences of *TP53*. The sequence files were aligned to human genome build 37 and 38 using bwa-mem2 (bwa v0.7.17) and processed via the GATK (v4.6.0.0) pipeline for variant discovery, excluding the step that removes duplicate reads to ensure maximum depth for low frequency variant detection. HaploTypeCaller (GATK) and LoFreq (v2.1.5) (56) were used to detect variants with an allele frequency of 1% or above in a sample. The clinical significance of a variant was determined by three primary criteria, 1) its frequency in the general population (gnomAD v4.0), 2) the submissions, classifications, and evidence reported to ClinVar, and 3) in-silico prediction algorithms. For missense variants, the likelihood of pathogenicity was evaluated by CADD (phred-scaled score >= 20)(57, 58), REVEL (score >= 0.5)(59), M-CAP (score >= 0.025)(60), DANN (score >= 0.96)(61), Bayes Del_ addAF(62, 63), AlphaMissense(64), GERP++(65), EVE(66) and EAp53(16, 67), EAp53 server is available at http://mammoth.bcm.tmc.edu/EAp53. Additionally, spliceAI(68) was implemented to predict potential splicing effects, with a score >= 0.3 as a determinant of higher likelihood. All variants were also classified as disruptive or non-disruptive according to Poeta et al. classification (15). These combined analyses allowed for a comprehensive in-silico evaluation to classify each variant. SNVs and indels were annotated using Ensembl Variant Effect Predictor (v112), based on the RefSeq transcript ID NM_000546.6 and Ensembl transcript ID ENST00000269305.9.

### Assessment of copy number alterations

Samples were genotyped with either the Global Diversity Array (v1.0, Illumina, Inc.), interrogating ∼1.8 million single nucleotide polymorphisms (SNPs) throughout the genome or the OmniExpressExome Array (v1.6, Illumina, Inc.), ∼962 thousand SNPs. The raw data was generated by the Illumina iScan system. Copy number alterations (CNAs) were detected with Nexus Copy Number (v10, Bionano, Inc) and MoChA (69). MoChA was also used to estimate the cell fraction of somatic chromosomal events. Low quality mosaic chromosomal alterations (mCAs) calls were excluded by implementing the following criteria: if the mCA length was greater than 2Mb, then the minimum values for lod_lrr_baf and lod_baf_phase were 10; if the mCA length was less than 2Mb, then the minimum value for lod_lrr_baf was 10 and the minimum value for lod_baf_phase was 30. Although MoChA finely distinguishes low frequency copy number neutral isodisomy events, it did not accurately quantify events with frequencies greater than 40%; high frequency CN-LOH events were either estimated to be less than 40% or not called at all. All samples were visually evaluated for somatic events using Nexus Copy Number, and high frequency mCAs observed but not detected by MoChA were quantified using a different method that implements a cumulative distribution function to evaluate a specific region of interest (70). For CNA analysis with Nexus Copy Number, the raw .idat files were processed with GenomeStudio (v2.0, Illumina, Inc.) using the GenCall software to call genotypes. SNPs with a GenTrain score < 0.7 were removed prior to downstream analysis. CNAs were detected by Nexus Copy Number using high performing SNPs with default parameters. For MoChA, the .idat files were converted to genotype call format (.gtc) using Illumina’s IAAP-CLI application, then merged via gtc2vcf (https://software.broadinstitute.org/software/gtc2vcf/).

### Outcomes and reporting of results

The primary outcome was defined as the detection of any variant with an allele frequency of 1% or above in the normal-appearing oral mucosa sample of a participant. Readouts for molecular testing were presented as the absence or presence of variants, the type of variant, description, allele frequency, frequency in the general population, predictions from in-silico algorithms, and classification of clinical significance according to ClinVar. As this is a research study and the significance of positive genetic findings from an oral brush biopsy collected from an area of normal mucosa is unknown, positive results were reported to patients’ physicians alone, to allow for closer surveillance. Participants were monitored for future clinical outcomes through their clinicians and pathology reports in case a biopsy was performed.

### Cytology and DNA ploidy analyses

Cells extracted from brushed lesions were subjected to clinical cytology testing and research DNA ploidy analysis, performed by the NCI/NIH, MD, US (NIH IRB protocol approval ID 001109 / CR001091) and the BC Cancer Research Institute, Vancouver, Canada (BCC, REB protocol approval number H22-02124), respectively. For cytology, samples were stained in accordance with the Papanicolaou method, interpreted by a certified cytopathologist and reported as negative, atypical, suspicious, positive, or indeterminate (13). DNA ploidy was assessed through DNA image cytometry to detect small percentages of aneuploid cells, a marker of malignancy and recorded as aneuploidy versus euploid (71). FA participants with oral lesions were consented to an NCI clinical trial (NCT05687149), and clinical cytology results were provided by the NIH.

### Droplet Digital PCR (ddPCR) for quantitation of FANCB duplication junction in gDNA

Copy number variance analysis was performed as was previously described (18) using the ddPCR system (Bio-Rad, Hercules, CA), adhering to the MIQE guidelines (72). The primers and FAM-labeled probe for the *FANCB* gDNA duplication junction were custom designed to target a 185 bp unique region with probe designed to hybridize to the *FANCB* exon 3-3 duplication junction in the gDNA of our samples. In the absence of the duplication junction in a given droplet, no PCR amplification of the *FANCB* exon 3-3 duplication product would take place. For the complete description of ddPCR method refer to Asur et al. (18).

### Statistical analysis

As a proof-of-concept study without hypotheses-driving cohorts’ size, we aimed to recruit at least 20 participants with FA and without FA, ensuring sufficient power (80%) to detect a 31% difference in the primary outcome - the presence of at least one molecular variant in the oral mucosa (α = 0.05). The non-FA cohort was used to establish the platform of keratinocyte extraction and DNA isolation in sufficient amounts and quality for the sequencing analysis of interest and to serve as control group. To match the younger age of FA patients, we analyzed results according to two age groups, below and above the age of 50 years. We assessed and compared baseline characteristics, as well as molecular results between the cohorts. Categorical variables were compared using a Chi-Square test or Fisher’s Exact Test. Continuous data were compared between two groups using a t-test or the Mann-Whitney test, as appropriate and between three groups by using nonparametric Kruskal-Wallis test with Dunn’s post hoc testing. A p-value of < 0.05 was used to denote statistical significance. Data was analyzed using SAS Studio software version 3.81 and Prism, version 10.

## Acknowledgments

We are indebted to the generosity of the FA patients and families, as well as to the volunteers who participated in this study. We thank the Smogorzewska lab members, the Rockefeller University Clinical Scholar program, facilitation office, hospital staff, and office of biostatistics, for their assistance. We acknowledge the support by the Fanconi Cancer Foundation (AS, CK, EV, NG, LM, DL, MG, SAS), Stand Up To Cancer-Fanconi Cancer Foundation-Farrah Fawcett Foundation Head and Neck Cancer Research Team Grant (AS), National Center for Advancing Translational Sciences, (UL1 TR001866) (TB and AS); the Shapiro-Silverberg Fund (TB); Stavros Niarchos Foundation (TB) and the Prevent Cancer Foundation (TB) and the Deutsche Fanconi-Anämie Hilfe e.V. (CK, EV). FXD, SS, ULH, KOA, JWT and SCC acknowledge support from the Intramural Research Program of the NIH National Human Genome Research Institute. The work of LJM, NG, AF and SAS was supported by the Intramural Research Program of the National Cancer Institute, National Institutes of Health. The work of ZK was supported by the Intramural Research Program of the National Institute of Dental and Craniofacial Research, National Institutes of Health. W.C. receives a fellowship related to this work from the Victorian Cancer Agency (MCRF21006). AI was used to proofread the manuscript.

## Authors Contribution

AS and TB conceived and designed this study and together with YCL consented all participants. TB and YCL aggregated cohort data from the IFAR, participants, and the treating physicians. TB collected majority of the brush samples. CK, EV, and FM collected some brush samples. TB and SS prepared DNA and TB, SS, KB, KOA, ULH, JWT collected sequencing and genotyping data. FXD, SCC, SS, KB, ULH, JWT, AS, and TB analyzed data. NG, LJM, ZK, SAS, and AF collected lesion samples and performed cytology analysis. DML and MG provided DNA image cytometry analysis. NG, LJM, ZK, EV, CK, WC, ADA, and RU provided clinical or sample information for subjects. TB performed biostatistical analysis of the data with the assistance of the Rockefeller University Office of Biostatistics. TB and AS wrote the manuscript with essential input from FXD and other authors. AS, TB, SCC, acquired funding. AS supervised the study.

## Data Availability Statement

All data are included in the Figures and Tables of the manuscript

## References

1. Johnson DE, Burtness B, Leemans CR, Lui VWY, Bauman JE, Grandis JR. Head and neck squamous cell carcinoma. Nat Rev Dis Primers. 2020;6(1):92.

2. Taylor AMR, Rothblum-Oviatt C, Ellis NA, Hickson ID, Meyer S, Crawford TO, et al. Chromosome instability syndromes. Nat Rev Dis Primers. 2019;5(1):64.

3. Kottemann MC, Smogorzewska A. Fanconi anaemia and the repair of Watson and Crick DNA crosslinks. Nature. 2013;493(7432):356–63.

4. Niraj J, Farkkila A, D’Andrea AD. The Fanconi Anemia Pathway in Cancer. Annu Rev Cancer Biol. 2019;3:457–78.

5. Smogorzewska A. The Fanconi Anemia DNA Repair Pathway. https://fanconi.org/clinical-care-guidelines/clinical-care/: Fanconi Cancer Foundation; 2020.

6. Kutler DI, Auerbach AD, Satagopan J, Giampietro PF, Batish SD, Huvos AG, et al. High incidence of head and neck squamous cell carcinoma in patients with Fanconi anemia. Arch Otolaryngol Head Neck Surg. 2003;129(1):106–12.

7. Kutler DI, Singh B, Satagopan J, Batish SD, Berwick M, Giampietro PF, et al. A 20-year perspective on the International Fanconi Anemia Registry (IFAR). Blood. 2003;101(4):1249–56.

8. Webster ALH, Sanders MA, Patel K, Dietrich R, Noonan RJ, Lach FP, et al. Genomic signature of Fanconi anaemia DNA repair pathway deficiency in cancer. Nature. 2022;612(7940):495–502.

9. Fiesco-Roa MO, Giri N, McReynolds LJ, Best AF, Alter BP. Genotype-phenotype associations in Fanconi anemia: A literature review. Blood Rev. 2019;37:100589.

10. Rosenberg PS, Socie G, Alter BP, Gluckman E. Risk of head and neck squamous cell cancer and death in patients with Fanconi anemia who did and did not receive transplants. Blood. 2005;105(1):67–73.

11. Scheckenbach K, Wagenmann M, Freund M, Schipper J, Hanenberg H. Squamous cell carcinomas of the head and neck in Fanconi anemia: risk, prevention, therapy, and the need for guidelines. Klin Padiatr. 2012;224(3):132–8.

12. Singh B. GJ, Kutler D. William W. Head and Neck Cancer in Patients with Fanconi Anemia. https://fanconi.org/clinical-care-guidelines/clinical-care/2020.

13. Velleuer E, Dietrich R, Pomjanski N, de Santana Almeida Araujo IK, Silva de Araujo BE, Sroka I, et al. Diagnostic accuracy of brush biopsy-based cytology for the early detection of oral cancer and precursors in Fanconi anemia. Cancer Cytopathol. 2020;128(6):403–13.

14. Lee RH, Kang H, Yom SS, Smogorzewska A, Johnson DE, Grandis JR. Treatment of Fanconi Anemia-Associated Head and Neck Cancer: Opportunities to Improve Outcomes. Clin Cancer Res. 2021;27(19):5168–87.

15. Poeta ML, Manola J, Goldwasser MA, Forastiere A, Benoit N, Califano JA, et al. TP53 mutations and survival in squamous-cell carcinoma of the head and neck. N Engl J Med. 2007;357(25):2552–61.

16. Neskey DM, Osman AA, Ow TJ, Katsonis P, McDonald T, Hicks SC, et al. Evolutionary Action Score of TP53 Identifies High-Risk Mutations Associated with Decreased Survival and Increased Distant Metastases in Head and Neck Cancer. Cancer Res. 2015;75(7):1527–36.

17. Wong YT, Tayeb MA, Stone TC, Lovat LB, Teschendorff AE, Iwasiow R, et al. A comparison of epithelial cell content of oral samples estimated using cytology and DNA methylation. Epigenetics. 2022;17(3):327–34.

18. Asur RS, Kimble DC, Lach FP, Jung M, Donovan FX, Kamat A, et al. Somatic mosaicism of an intragenic FANCB duplication in both fibroblast and peripheral blood cells observed in a Fanconi anemia patient leads to milder phenotype. Mol Genet Genomic Med. 2018;6(1):77–91.

19. Smetsers SE, Velleuer E, Dietrich R, Wu T, Brink A, Buijze M, et al. Noninvasive molecular screening for oral precancer in Fanconi anemia patients. Cancer Prev Res (Phila). 2015;8(11):1102–11.

20. Poell JB, Wils LJ, Brink A, Dietrich R, Krieg C, Velleuer E, et al. Oral cancer prediction by noninvasive genetic screening. Int J Cancer. 2023;152(2):227–38.

21. Errazquin R, Carrasco E, Del Marro S, Sunol A, Peral J, Ortiz J, et al. Early Diagnosis of Oral Cancer and Lesions in Fanconi Anemia Patients: A Prospective and Longitudinal Study Using Saliva and Plasma. Cancers (Basel). 2023;15(6).

22. Endler G, Greinix H, Winkler K, Mitterbauer G, Mannhalter C. Genetic fingerprinting in mouthwashes of patients after allogeneic bone marrow transplantation. Bone Marrow Transplant. 1999;24(1):95–8.

23. Thiede C, Prange-Krex G, Freiberg-Richter J, Bornhauser M, Ehninger G. Buccal swabs but not mouthwash samples can be used to obtain pretransplant DNA fingerprints from recipients of allogeneic bone marrow transplants. Bone Marrow Transplant. 2000;25(5):575–7.

24. Garaycoechea JI, Crossan GP, Langevin F, Mulderrig L, Louzada S, Yang F, et al. Alcohol and endogenous aldehydes damage chromosomes and mutate stem cells. Nature. 2018;553(7687):171–7.

25. Garaycoechea JI, Crossan GP, Langevin F, Daly M, Arends MJ, Patel KJ. Genotoxic consequences of endogenous aldehydes on mouse haematopoietic stem cell function. Nature. 2012;489(7417):571–5.

26. Langevin F, Crossan GP, Rosado IV, Arends MJ, Patel KJ. Fancd2 counteracts the toxic effects of naturally produced aldehydes in mice. Nature. 2011;475(7354):53–8.

27. Dingler FA, Wang M, Mu A, Millington CL, Oberbeck N, Watcham S, et al. Two Aldehyde Clearance Systems Are Essential to Prevent Lethal Formaldehyde Accumulation in Mice and Humans. Mol Cell. 2020;80(6):996–1012 e9.

28. Pontel LB, Rosado IV, Burgos-Barragan G, Garaycoechea JI, Yu R, Arends MJ, et al. Endogenous Formaldehyde Is a Hematopoietic Stem Cell Genotoxin and Metabolic Carcinogen. Mol Cell. 2015;60(1):177–88.

29. Wang M, Brandt LTL, Wang X, Russell H, Mitchell E, Kamimae-Lanning AN, et al. Genotoxic aldehyde stress prematurely ages hematopoietic stem cells in a p53-driven manner. Mol Cell. 2023;83(14):2417–33 e7.

30. Mu A, Hira A, Mori M, Okamoto Y, Takata M. Fanconi anemia and Aldehyde Degradation Deficiency Syndrome: Metabolism and DNA repair protect the genome and hematopoiesis from endogenous DNA damage. DNA Repair (Amst). 2023;130:103546.

31. Hira A, Yabe H, Yoshida K, Okuno Y, Shiraishi Y, Chiba K, et al. Variant ALDH2 is associated with accelerated progression of bone marrow failure in Japanese Fanconi anemia patients. Blood. 2013;122(18):3206–9.

32. Jung M, Kim J, Park Y, Ilyashov I, Yang F, Choijilsuren HB, et al. ALDH9A1 deficiency as a source of endogenous DNA damage that requires repair by the Fanconi anemia pathway. J Cell Biol. 2025;224(7).

33. Blobel NJ, Yao Y, Okondo MC, Smogorzewska A. Genotoxic formaldehyde and lipid aldehydes are sources of DNA damage in keratinocytes. bioRxiv. 2025.

34. Jaiswal S, Fontanillas P, Flannick J, Manning A, Grauman PV, Mar BG, et al. Age-related clonal hematopoiesis associated with adverse outcomes. N Engl J Med. 2014;371(26):2488–98.

35. Genovese G, Kahler AK, Handsaker RE, Lindberg J, Rose SA, Bakhoum SF, et al. Clonal hematopoiesis and blood-cancer risk inferred from blood DNA sequence. N Engl J Med. 2014;371(26):2477–87.

36. Martincorena I, Roshan A, Gerstung M, Ellis P, Van Loo P, McLaren S, et al. Tumor evolution. High burden and pervasive positive selection of somatic mutations in normal human skin. Science. 2015;348(6237):880–6.

37. Martincorena I, Campbell PJ. Somatic mutation in cancer and normal cells. Science. 2015;349(6255):1483–9.

38. Martincorena I, Fowler JC, Wabik A, Lawson ARJ, Abascal F, Hall MWJ, et al. Somatic mutant clones colonize the human esophagus with age. Science. 2018;362(6417):911–7.

39. Lee-Six H, Olafsson S, Ellis P, Osborne RJ, Sanders MA, Moore L, et al. The landscape of somatic mutation in normal colorectal epithelial cells. Nature. 2019;574(7779):532–7.

40. Fiala C, Diamandis EP. Mutations in normal tissues-some diagnostic and clinical implications. BMC Med. 2020;18(1):283.

41. Yuan DJ, Zinno J, Botella T, Dhingra D, Wang S, Hawkins A, et al. Genotype-to-phenotype mapping of somatic clonal mosaicism via single-cell co-capture of DNA mutations and mRNA transcripts. bioRxiv. 2024.

42. Yizhak K, Aguet F, Kim J, Hess JM, Kubler K, Grimsby J, et al. RNA sequence analysis reveals macroscopic somatic clonal expansion across normal tissues. Science. 2019;364(6444).

43. Lawson ARJ, Abascal F, Nicola PA, Lensing SV, Roberts AL, Kalantzis G, et al. Somatic mutation and selection at population scale. Nature. 2025;647(8089):411–20.

44. Chabanon RM, Danlos FX, Ouali K, Postel-Vinay S. Genome instability and crosstalk with the immune response. Genome Med. 2025;17(1):139.

45. Curtis RE, Rowlings PA, Deeg HJ, Shriner DA, Socie G, Travis LB, et al. Solid cancers after bone marrow transplantation. N Engl J Med. 1997;336(13):897–904.

46. Leemans CR, Braakhuis BJ, Brakenhoff RH. The molecular biology of head and neck cancer. Nat Rev Cancer. 2011;11(1):9–22.

47. Curtis RE, Metayer C, Rizzo JD, Socie G, Sobocinski KA, Flowers ME, et al. Impact of chronic GVHD therapy on the development of squamous-cell cancers after hematopoietic stem-cell transplantation: an international case-control study. Blood. 2005;105(10):3802–11.

48. Kim SY, Kim GJ, Bang JI, Shin HI, Sun DI. Are second primary head and neck cancers with previous hematological malignancy more aggressive than de novo head and neck cancers? Am J Otolaryngol. 2023;44(2):103748.

49. Shimada K, Yokozawa T, Atsuta Y, Kohno A, Maruyama F, Yano K, et al. Solid tumors after hematopoietic stem cell transplantation in Japan: incidence, risk factors and prognosis. Bone Marrow Transplant. 2005;36(2):115–21.

50. Witherspoon RP, Fisher LD, Schoch G, Martin P, Sullivan KM, Sanders J, et al. Secondary cancers after bone marrow transplantation for leukemia or aplastic anemia. N Engl J Med. 1989;321(12):784–9.

51. Ramirez MJ, Pujol R, Trujillo-Quintero JP, Minguillon J, Bogliolo M, Rio P, et al. Natural gene therapy by reverse mosaicism leads to improved hematology in Fanconi anemia patients. Am J Hematol. 2021;96(8):989–99.

52. Nicoletti E, Rao G, Bueren JA, Rio P, Navarro S, Surralles J, et al. Mosaicism in Fanconi anemia: concise review and evaluation of published cases with focus on clinical course of blood count normalization. Ann Hematol. 2020;99(5):913–24.

53. Rio P, Navarro S, Wang W, Sanchez-Dominguez R, Pujol RM, Segovia JC, et al. Successful engraftment of gene-corrected hematopoietic stem cells in non-conditioned patients with Fanconi anemia. Nat Med. 2019;25(9):1396–401.

54. Gutkind JS, Molinolo AA, Wu X, Wang Z, Nachmanson D, Harismendy O, et al. Inhibition of mTOR signaling and clinical activity of metformin in oral premalignant lesions. JCI Insight. 2021;6(17).

55. Harris PA, Taylor R, Thielke R, Payne J, Gonzalez N, Conde JG. Research electronic data capture (REDCap)--a metadata-driven methodology and workflow process for providing translational research informatics support. J Biomed Inform. 2009;42(2):377–81.

56. Wilm A, Aw PP, Bertrand D, Yeo GH, Ong SH, Wong CH, et al. LoFreq: a sequence-quality aware, ultra-sensitive variant caller for uncovering cell-population heterogeneity from high-throughput sequencing datasets. Nucleic Acids Res. 2012;40(22):11189–201.

57. Kircher M, Witten DM, Jain P, O’Roak BJ, Cooper GM, Shendure J. A general framework for estimating the relative pathogenicity of human genetic variants. Nat Genet. 2014;46(3):310–5.

58. Schubach M, Maass T, Nazaretyan L, Roner S, Kircher M. CADD v1.7: using protein language models, regulatory CNNs and other nucleotide-level scores to improve genome-wide variant predictions. Nucleic Acids Res. 2024;52(D1):D1143–D54.

59. Hopkins JJ, Wakeling MN, Johnson MB, Flanagan SE, Laver TW. REVEL Is Better at Predicting Pathogenicity of Loss-of-Function than Gain-of-Function Variants. Hum Mutat. 2023;2023:8857940.

60. Jagadeesh KA, Wenger AM, Berger MJ, Guturu H, Stenson PD, Cooper DN, et al. M-CAP eliminates a majority of variants of uncertain significance in clinical exomes at high sensitivity. Nat Genet. 2016;48(12):1581–6.

61. Quang D, Chen Y, Xie X. DANN: a deep learning approach for annotating the pathogenicity of genetic variants. Bioinformatics. 2015;31(5):761–3.

62. Feng BJ. PERCH: A Unified Framework for Disease Gene Prioritization. Hum Mutat. 2017;38(3):243–51.

63. Tian Y, Pesaran T, Chamberlin A, Fenwick RB, Li S, Gau CL, et al. REVEL and BayesDel outperform other in silico meta-predictors for clinical variant classification. Sci Rep. 2019;9(1):12752.

64. Cheng J, Novati G, Pan J, Bycroft C, Zemgulyte A, Applebaum T, et al. Accurate proteome-wide missense variant effect prediction with AlphaMissense. Science. 2023;381(6664):eadg7492.

65. Davydov EV, Goode DL, Sirota M, Cooper GM, Sidow A, Batzoglou S. Identifying a high fraction of the human genome to be under selective constraint using GERP++. PLoS Comput Biol. 2010;6(12):e1001025.

66. Frazer J, Notin P, Dias M, Gomez A, Min JK, Brock K, et al. Disease variant prediction with deep generative models of evolutionary data. Nature. 2021;599(7883):91–5.

67. Michikawa C, Torres-Saavedra PA, Silver NL, Harari PM, Kies MS, Rosenthal DI, et al. Evolutionary Action Score of TP53 Analysis in Pathologically High-Risk Human Papillomavirus-Negative Head and Neck Cancer From a Phase 2 Clinical Trial: NRG Oncology Radiation Therapy Oncology Group 0234. Adv Radiat Oncol. 2022;7(6):100989.

68. Jaganathan K, Kyriazopoulou Panagiotopoulou S, McRae JF, Darbandi SF, Knowles D, Li YI, et al. Predicting Splicing from Primary Sequence with Deep Learning. Cell. 2019;176(3):535–48 e24.

69. Loh PR, Genovese G, Handsaker RE, Finucane HK, Reshef YA, Palamara PF, et al. Insights into clonal haematopoiesis from 8,342 mosaic chromosomal alterations. Nature. 2018;559(7714):350–5.

70. Markello TC, Carlson-Donohoe H, Sincan M, Adams D, Bodine DM, Farrar JE, et al. Sensitive quantification of mosaicism using high density SNP arrays and the cumulative distribution function. Mol Genet Metab. 2012;105(4):665–71.

71. Datta M, Laronde DM, Rosin MP, Zhang L, Chan B, Guillaud M. Predicting Progression of Low-Grade Oral Dysplasia Using Brushing-Based DNA Ploidy and Chromatin Organization Analysis. Cancer Prev Res (Phila). 2021;14(12):1111–8.

72. Huggett JF, Foy CA, Benes V, Emslie K, Garson JA, Haynes R, et al. The digital MIQE guidelines: Minimum Information for Publication of Quantitative Digital PCR Experiments. Clin Chem. 2013;59(6):892–902.

