## Supplemental data for "Fanconi Anemia as a Window into Premalignant Field Cancerization of the Oral Mucosa"

**Supplemental materials include:**
Figure S1
Table S1-S3
Table S4 (excel)
Supplemental methods

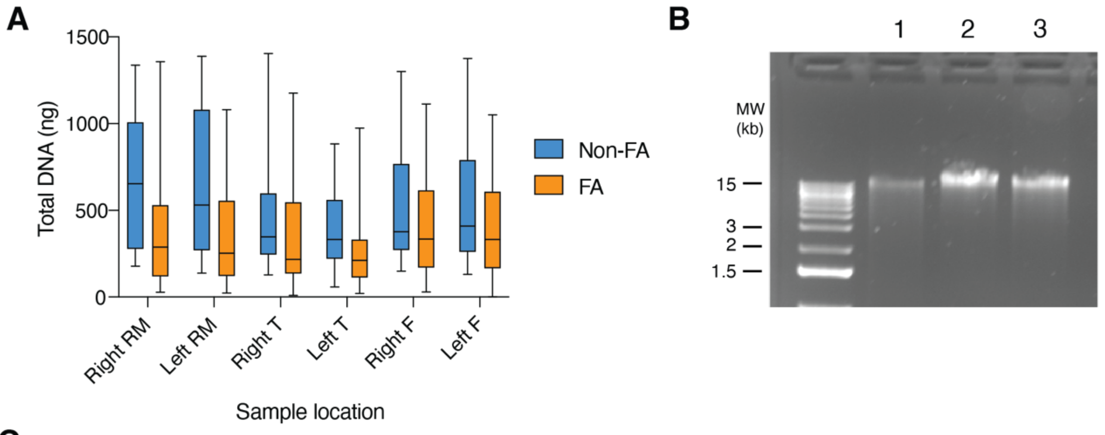

**C**

| **Brushed area** | **Non-FA controls (n=35), median DNA amount, ng (IQR)** | **FA (n=38), median DNA amount, ng (IQR)** |
|---|---|---|
| Retromolar area | 588.25 (270-980.3) | 276.1 (120.4-530) |
| Lateral tongue | 336.15 (231.5-589.8) | 212.3 (120.6-530) |
| Floor of the mouth | 379.4 (268.8-764.1) | 331.8 (168.8-599.6) |

**Supplementary Figure 1**. **DNA yield per oral biopsy site brushed, among the two cohorts.** (**A**) Box and whisker plot depicting the distribution of DNA amounts in nanograms per each of the six normal appearing oral mucosa sites brushed for the healthy control cohort (HC) and Fanconi anemia cohort (FA). The middle line inside the box denotes the median DNA amount, while the whiskers represent the range of the data, from the minimum to quartile 1 (Q1) and from quartile 3 (Q3) to the maximum. Statistical comparison between cohorts was not conducted as most of the Fanconi anemia cohort was brushed with 2 brushes per site to allow for enough DNA per site, while half of the non-FA control cohort was brushed using a single brush. (**B**) Ethidium bromide (EtBr) gel electrophoresis of three oral brush samples, collected from the lateral tongue of two healthy controls, showing high molecular weight DNA. DNA concentration for these DNA samples was 12, 22.2 and 22.7 ng/ul, for samples 1, 2, and 3, respectively. (**C**) Quantification of median DNA yield and interquartile range (Q1-Q3) per each of the anatomical sites brushed, for the two cohorts. RM – retromolar; T – lateral tongue ; F – floor of the mouth. HC – healthy control cohort; FA – Fanconi anemia cohort; IQR – interquartile range

**Supplementary Table 1.** List of somatic copy number events found in oral brushes from normal appearing mucosa and saliva samples of Fanconi anemia patients. Variants perceived as germline, according to their presence and even distribution throughout the six normal appearing sites sampled or their presence in the blood or skin sample, are not included. "Estimated cell fraction" is the estimated proportion of cells harboring an indicated alteration.

| | | Somatic copy number alterations (CNA) event | | | |
|---|---|---|---|---|---|
| **FA participant number** | **Biopsy location** | **Chr** | **Position (GRCh37)** | **Type of event** | **Estimated cell fraction** |
| 24 | Right lateral tongue | 3p | 70,374,871 – 83,465,789 | deletion | 20% |
| | | 9p | 1 – 40,804,094 | isodisomy | 19% |
| | | 17q | 39,248,333 – 81,195,210 | isodisomy | 6% |
| 25 | Right retromolar area | 9p | 1 - 34,971,963 | isodisomy | 33% |
| | | 9p | 35,177,345 - 38,254,018 | isodisomy | 18% |
| | | 16q | 83,925,476 - terminus | isodisomy | 4% |
| | Right lateral tongue | 9p | 1 - 36,572,757 | isodisomy | 35% |
| | | 4p | 1 - 10,363,994 | deletion | 24% |
| | | 16q | 87,714,039 – terminus | deletion | 23% |
| | | 17q | 40,222,638 - 81,195,210 | isodisomy | 14% |
| | Pooled sample: left lateral tongue + floor of the mouth | 9p | 1 - 36,349,561 | isodisomy | 12% |
| | | 16q | 49,630,762 - 61,796,595 | isodisomy | 7% |
| | | 16q | 62,025,800 - 66,386,711 | isodisomy | 20% |
| | | 16q | 66,391,517 - 83,912,536 | isodisomy | 22% |
| | | 16q | 83,925,476 - terminus | isodisomy | 25% |
| 26 | Right side of the floor of mouth | 9p | 1 - 16,918,196 | isodisomy | 4% |
| | Right lateral tongue | 9p | 1 - 26,455,313 | isodisomy | 7% |
| | Left lateral tongue | 9p | 1 - 21,991,923 | isodisomy | 10% |
| | | 9p | 22,412,570 - 33,430,585 | isodisomy | 10% |
| 27 | Right lateral tongue | 9p | 1 - 36,815,627 | isodisomy | 8% |

| | | | | | |
|---|---|---|---|---|---|
| | Pooled sample: Left retromolar area + Right side of the floor of mouth + Left side of the floor of mouth | 9p | 1 - 36,815,627 | isodisomy | 34% |
| | | 8q | 87,694,185 - 97,172,671 | deletion | 38% |
| | | 6 | entire chromosome | duplication | 21% |
| 28 | Right retromolar area | 15q | 45,404,066 - terminus | isodisomy | 20% |
| | | 22q | 19,924,021 - terminus | isodisomy | 13% |
| | Left retromolar area | 15q | 45,404,066 - terminus | isodisomy | 24% |
| | | 22q | 18,941,948 - terminus | isodisomy | 6% |
| | Right side of the tongue | 15q | 40,177,769 - terminus | isodisomy | 22% |
| | | 22q | 26,132,612 - terminus | isodisomy | 4% |
| | Left side of the tongue | 15q | 40,177,769 - terminus | isodisomy | 22% |
| | | 22q | 27,661,009 - terminus | isodisomy | 4% |
| | Right side of the floor of mouth | 15q | 24,250,979 - terminus | isodisomy | 31% |
| | | 22q | 16,000,000 - terminus | isodisomy | 7% |
| | Left side of the floor of mouth | 15q | 41,600,878 - terminus | isodisomy | 27% |
| | | 22q | 19,889,423 - terminus | isodisomy | 14% |
| | Saliva | 15q | 89,324,025 - 102,531,392 | isodisomy | 30% |
| | | 22q | 19,766,782 - 51,304,566 | isodisomy | 80% |
| 29 | Right lateral tongue | 6q | 134,251,090 - 171,115,067 | deletion | 15% |
| | | 17p | 45,933,165 – 81,195,210 | duplication | 16% |
| | Right side of the floor of mouth | 13q | 57,306,274 - 70,507,227 | deletion | 12% |
| 32 | Left retromolar area | 1q | 170,152,927 – terminus | duplication | 17% |
| | | 1q | 197,266,535 - 240,805,113 | duplication | 42% |
| | | 10q | 133,129,073 – terminus | deletion | 25% |
| | | 15q | 25,721,212 – terminus | isodisomy | 22% |
| | | 17p | 4,075,158 - 5,437,285 | deletion | 26% |
| | Right lateral tongue | 6p | 2,774,052 - 47,065,653 | isodisomy | 49% |
| | Right side of the floor of mouth | Many Chromosomal mosaic events (refer to supplementary table 5) | | | |

| | | | | | |
|---|---|---|---|---|---|
| | Left side of the floor of mouth | Many chromosomal mosaic events similar to the right side of the floor of the mouth, with a lower allele frequency of events | | | |
| 33 | Right lateral tongue | 3q | 94,786,646 - 125,786,760<br>125,798,083 - 144,340,205<br>144,342,672 - 149,911,847<br>149,974,934 - 150,398,254<br>150,398,288 - 151,509,744<br>151,554,749 - 151,887,185 | gain | 11%<br>15%<br>24%<br>76%<br>41%<br>47% |
| | | 6p | 30,805,921 - 62,694,479 | isodisomy | 7% |
| | | 7q | 55,330,141 - 159,138,663 | isodisomy | 7% |
| | | 17p | 1 - 8,613,462 | isodisomy | 10% |
| | | 17p | 8,614,578-17,567,943 | isodisomy | 8% |
| | | 19p | 0 - 20906994 | isodisomy | 10% |
| | Saliva | 6q | 168,342,746 - 168,595,832 | gain | 76% |
| 34 | Right lateral tongue | 4q | 184,060,896 – 188,980,585 | deletion | 23% |
| | | 9p | 1 – 21,944,627 | isodisomy | 19% |
| | | 9p | 22,017,550 – 32,579,732 | isodisomy | 26% |
| | | 9p | 32,701,164 – 36,872,314 | isodisomy | 19% |
| | Left lateral tongue | 7 | entire chromosome | gain | 23% |
| | Left side of the floor of the mouth | 9p | 1 – 34,918,201 | isodisomy | 9% |
| | Saliva | 1q | 144,549,763 - 249,250,621 | gain | 13% |
| 36 | Left lateral tongue | 1q | 156,063,090-249,250,621 | gain | 32% |
| | | 8 | entire chromosome | gain (trisomy 8) | 36% |
| 37 | Right retromolar area | 3p | 1 – 54,202,453 | isodisomy | 12% |
| | Right lateral tongue | 3p | 1 – 51,812,952 | isodisomy | 9% |

| | | | | | |
|---|---|---|---|---|---|
| | Right side of the floor of the mouth | 3p | 1 – 29,700,225 | isodisomy | 26% |
| | | 3q | 29,707,118 - 48,327,573 | isodisomy | 22% |
| | Left side of the floor of the mouth | 3p | 1 - 13,053,744 | isodisomy | 25% |
| | | 3p | 13,053,937 - 45,226,973 | isodisomy | 15% |
| | | 3p | 45,237,733 - 63,470,819 | isodisomy | 9% |
| 41 | Right retromolar area | 9p | 1 - 47202144 | isodisomy | 90-100% |
| | Left buccal area | 9p | 1 - 37503087 | isodisomy | 6% |
| | Right lateral tongue | 9p | 1 - 32061188 | isodisomy | 10% |
| | | 22q | 44799094 - terminus | isodisomy | 10% |
| | Left lateral tongue | 9p | 1 - 35826038 | isodisomy | 25% |
| | | 22q | 25602790 - terminus | isodisomy | 29% |
| | Right side of the floor of mouth | 9p | 1 - 32082616 | isodisomy | 5% |
| | Left side of the floor of mouth | 9p | 1 - 35333961 | isodisomy | 20% |
| | | 22q | 31233875 - 51304566 | isodisomy | 10% |
| 42 | Right lateral tongue | 9p | 1 - 32218797 | isodisomy | 5% |
| | | 17q | 55954403-81195210 | gain | 23% |
| | | 20q | 55856403 - 63025520 | isodisomy | 9% |
| 67 | Left retromolar area | 1q | 200,300,768 - 249,212,557 | duplication | ~30% |
| | | 9p | 1 - 47,202,144 | isodisomy | ~35% |
| 69 | Left retromolar area | 9p | 1-34513110 | isodisomy | ~75% |
| | | 9p | 21496058-22697407 | deletion | ~75% |
| | Left lateral tongue | 1q | 142550999-189354243 | isodisomy | ~30% |
| | | 1q | 236407467- 249212725 | isodisomy | ~30% |
| | | Xp | 4714113- 45332458 | isodisomy | ~30% |
| 71 | Right side of the floor of the mouth | 1p | 0 – 21,848,962 | isodisomy | 14% |
| 75 | Left lateral tongue | 1q | 142,544,928-239,192,955 | trisomy | 15% |
| | | 9p | 1-47,202,144 | isodisomy | 50% |
| 76 | Right lateral tongue | 9p | 1 – 45,377,977 | isodisomy | 95% |

| | | | | | |
|---|---|---|---|---|---|
| | Left lateral tongue | 6q | 75,980,587 - 166,013,760 | isodisomy | 35% |
| | | 8 | 1 - 146,364,022 (entire chromosome) | gain | 22% |
| | Right side of the floor of mouth | 9p | 1 – 40,804,094 | isodisomy | 8% |
| | Left side of the floor of mouth | 11q | 111,470,567 - 135,006,516 | deletion | 12% |
| 78 | Right lateral tongue | 1q | 173,686,353 - 249,250,621 | gain | 17% |
| | | 8q | 46,900,284 - 137,688,230 | gain | 40% |
| | | 8q | 137,865,786 - 146,364,022 | gain | 42% |
| | Left side of the floor of mouth | 3p | 1 – 48,680,470 | isodisomy | 35% |
| 79 | Right retromolar area | 22q | 16,000,000 - 28,121,605 | isodisomy | 8% |
| | | 22q | 28,128,858 - 51,304,566 | isodisomy | 10% |
| | Left retromolar area | 22q | 18,628,212 - 51,304,566 | isodisomy | 12% |
| | Right lateral tongue | 22q | 16,000,000,- 18,320,886 | isodisomy | 11% |
| | | 22q | 18,323,438 - 30,557,278 | isodisomy | 19% |
| | | 22q | 30,597,810 - 51,304,566 | isodisomy | 22% |
| | Left lateral tongue | 22q | 19,639,383 - 27,717,833 | isodisomy | 20% |
| | | 22q | 27,718,617 - 51,304,566 | isodisomy | 25% |
| | Right side of the floor of the mouth | 22q | 18,323,438 - 24,550,009 | isodisomy | 10% |
| | | 22q | 24,622,648 - 51,304,566 | isodisomy | 15% |
| | Left side of the floor of the mouth | 22q | 19,675,944 - 51,304,566 | isodisomy | 8% |
| 80 | Left retromolar area | 9p | 0 – 37,686,792 | isodisomy | 80% |
| | Right side of the tongue | 1q | 175,210,458 - 249,250,621 | gain | 20% |
| | | 4q | 178,340,126 - 191,154,276 | deletion | 23% |
| | Left side of the tongue | 21q | 37,744,825 - 37,935,548 | deletion | 39% |
| | | 21q | 41,792,388 - 47,606,248 | deletion | 35% |
| 81 | Right retromolar area | 14q | 21,549,893 - 107,289,356 | isodisomy | 35% |
| | Left retromolar area | 14q | 20,975,984 – 23,094,197 | isodisomy | 9% |
| | | 14q | 23,094,197 – 107,289,356 | isodisomy | 45% |
| | | 9p | 1 – 33,078,031 | isodisomy | 15% |
| | Right lateral tongue | 9p | 1 – 37,944,489 | isodisomy | 15% |
| | | 14q | 20,975,984 – 23,094,197 | isodisomy | 15% |

| | | | | | |
|---|---|---|---|---|---|
| | Left lateral tongue | 14q | 23,094,197 - 107,289,356 | isodisomy | 75% |
| | Right side of the floor of mouth | 14q | 23,094,197 - 107,289,356 | isodisomy | 30% |
| | Left side of the floor of mouth | 14q | 23,094,197 - 107,289,356 | isodisomy | 30% |
| | Saliva | 14q | 37,311,528 - 107,289,356 | isodisomy | 100% |
| 83 | Saliva | 1q | 146,501,334 - 249,250,621 | gain | 23% |
| | | 11q | 103,032,428 - 135,006,516 | deletion | 26% |
| 85 | Left side of the floor of the mouth | 6q | 72,451,179 - 124,951,063 | undetermined | 10-30% |

**Supplementary Table 2. A detailed list of *TP53* variants found in oral lesions from Fanconi anemia individuals and their corresponding annotation.** Table describes the oral location of variants found, variants' allele frequency, location on chromosome 17 (Chr.17) and the exon number, the nucleotide in the reference sequence (Ref) and the alteration found (Alt), the type of functional genetic variants, the notation of the specific genetic variant in the complementary DNA (cDNA) and the protein level change results from the genetic variant in the gene, the frequency of the variants in the general population according to gnomAD v.4, its classifications in ClinVar with number of submissions and in silico prediction scores of variant's effect using various computational algorithms.

| FA # | Location | Allele frequency | Position on Chr.17 GRCh37 | Exon | Ref | Alt | Variant type | cDNA | Protein | General population frequency (gnomAD v4.0) | ClinVar - number and type of submissions at time of analysis | | Variant effect prediction scores | | | | | | | | | |
|---|---|---|---|---|---|---|---|---|---|---|---|---|---|---|---|---|---|---|---|---|---|---|
| | | | | | | | | | | | | | CADD v1.7 | DANN | BayesDel_addAF | REVEL | AlphaMissense | SpliceAI | GERP++ | EVE | Poeta et al. | EAp53 |
| 32 | Leuko/ erythroplakia at the left lateral-tip of the tongue | 1% | 7576855 | 9 | G | A | stop gain | c.991C>T | p.Q331* | 0 | 5 | P | 38 | 0.977 | D | -- | -- | -- | 2.88 | -- | Dis | -- |
| | | 12% | 7577121 | 8 | G | A | missense | c.817C>T | p.R273C | 6/1613804 | Multiple | P / LP | 27.1 | 0.999 | D | 0.901 | LP | -- | 3.95 | P | ND | 84.52 High |
| 41 | Ulcer at the left lateral tongue | 9% | 7578268 | 6 | A | C | missense | c.581T>G | p.L194R | 1/1614188 | 5 | P / LP / US | 26.8 | 0.997 | D | 0.933 | LP | -- | 5.41 | P | Dis | 91.12 High |
| | Ulcer at the left lateral tongue<br>Erythroplakia right upper gingiva,<br>Erythroplakia left upper gingiva<br>Saliva sample | 1%<br>1%<br>1%<br>3% | 7578393 | 5 | A | T | missense | c.537T>A | p.H179Q | 0 | ** | P | 13.41 | 0.980 | D | 0.79 | LP | -- | -9.19 | | Dis | 74.44 Low |
| 69 | Right lateral tongue lesion | 1.1% | 7577509 | 7 | C | A | stop gain | c.772G>T | p.E258* | 0 | 4 | P | 49 | 0.995 | D | -- | -- | -- | 3.62 | -- | Dis | -- |
| | | 3.2% | 7579412-7579419 | 4 | GGCCAGGA | - | indel | c.268_275 del | p.S90Pfs*56 | 0 | 0 | None | -- | -- | -- | -- | -- | -- | -- | -- | Dis | -- |
| 80 | Top right anterior tongue lesion | 12% | 7574018 | 10 | G | A | missense | c.1009C>T | p.R337C | 0 | ** | P | 22.1 | 0.97 | D | 0.715 | P | -- | 3.38 | B | ND | 63.66 Low |

** ClinVar classifies this variant based on a submission by the ClinGen *TP53* Variation Curation Expert Panel, a FDA recognized database. Lt.–left; Rt.–right; SNV – single nucleotide variant; P – pathogenic; LP- likely pathogenic; US- uncertain significance; CADD - Combined Annotation Dependent Depletion; DANN - Deep Artificial Neural Network; BayesDel_addAF - Bayesian Deleteriousness model with added Allele Frequency; REVEL - Rare Exome Variant Ensemble Learner; SpliceAI - Splicing Artificial Intelligence; GERP - Genomic Evolutionary Rate Profiling; EVE - Evolutionary model of Variant Effect; D – Deleterious; Dis – Disruptive or ND – nondisruptive mutation (15); EAp53 - evolutionary action of *TP53* scoring algorithm – scores missense mutations from 0 to 100, with higher (H) scores indicate more deleterious effects than lower (L) scores (16)

**Supplementary Table 3.** List of somatic copy number events found in oral brushes collected from lesions of Fanconi anemia patients.

| FA participant number | Biopsy location | Somatic copy number alterations (CNA) event | | | |
|---|---|---|---|---|---|
| | | Chr | Position (GRCh37) | Type of event | Estimated cell fraction |
| 32 | Leuko/erythroplakia at the left lateral-tip of the tongue | 1q | 190,174,840 - 201,159,461 | duplication | 17% |
| | | 1q | 201,174,204 - 248,401,116 | duplication | 24% |
| | Leukoplakia located to the right of the tip of the tongue | 1q | 25,721,212-102,531,392 | isodisomy | 19% |
| | | 3p | 1 - 94,452,002 | deletion | 21% |
| | | 3q | 94,455,418 - 181,681,668 | duplication | 15% |
| | | 3q | 187,685,110 - 198,022,430 | duplication | 20% |
| | | 9p | 1 - 37,277,603 | isodisomy | 19% |
| | | 17p | 1 - 16,824,504 | deletion | 22% |
| | Gingival lesion at the right lower jaw | Many chromosomal mosaic events similar to those found at the right and left side of the floor of the mouth, with a lower allele frequency of events (see supplementary table 5) | | | |
| 37 | Lesion at the left lateral tongue | 3p | 1 - 47,242,923 | isodisomy | 7% |
| | Lesion at the right lateral tongue | 3p | 1 - 30,775,466 | isodisomy | 8% |
| 41 | Ulcer at the left lateral tongue | 2q | 176,738,540 - terminus | isodisomy | 10% |
| | | 8p | 1 - 27,474,202 | isodisomy | 11% |
| | | 9p | 1 - 31,146,393 | isodisomy | 12% |
| 79 | Lesion at the top of the tongue | 22q | 18,618,827 - 51,304,566 | isodisomy | 15% |
| 80 | Lesion at the top right anterior area of the tongue | 3p | 1 - 90,307,685 | deletion | 18% |
| | | 9p | 293,35,633 - 33,007,040 | gain | 35% |
| | | 13q | 19,000,000 - 27,099,528 | deletion | 19% |
| | | 13q | 27,100,247 - 115,169,878 | deletion | 25% |
| | | 17p | 1 - 21,901,755 | deletion | 27% |
| | | 17q | 27,382,061 - 46,799,812 | deletion | 18% |
| 81 | Lesion at the right border of the lateral/tip of tongue | 14q | 20,975,984 – 23,094,197 | isodisomy | 10% |
| | | 14q | 23,094,197 – 107,289,356 | isodisomy | 65% |
| 85 | Lesion at the right lateral tongue | 1q ter | 171,610,484 - 249,212,725 | duplication | 25-50% |

### Supplemental methods

#### Oral brushes and DNA isolation

Noninvasive oral brush biopsy samples were stored in 'SurePath' preservation solution. DNA isolation failed for the samples processed from the first two healthy controls. Using an optimized DNA isolation protocol, we were able to extract oral nucleated keratinocytes and obtain enough high molecular weight DNA for sequencing analysis, from each of the six distinct regions of normal appearing mucosa. Both freshly collected and stored samples were successfully used for molecular studies. Storing of non-FA volunteer samples in SurePath preservation solution, for an average duration of 21 days (range 0-77 days), did not impair DNA yield or sequencing data. Majority of FA oral brush samples were processed either fresh or within days, with an average storage time of 9 days (range 0-70 days). DNA yield varied among participants and the three different anatomical sites brushed, without a uniform trend (**Supplementary Figure 1**). Cells quantification of samples had a poor correlation with total DNA yield. We did not observe a pattern of consistent decay of DNA in correlation to the length of samples' storage (**data not shown**). Median DNA quantities isolated from 108 brushed samples of the first 18 healthy recruited participants was 262.8ng (interquartile range, [IQR] 198.9-326.8ng). In comparison to the HC cohort, oral brush samples collected from the first five FA individuals yielded lower DNA amounts (median 114ng, IQR1-3 of 52.3-155.2ng), resulting in a protocol change to using two oral brushes per site, instead of one. With this amendment, median DNA quantities increased to 471.2ng (IQR 1-3 of 233.6-757.6ng).
